# Evaluation of patient-specific cell free DNA assays for monitoring of minimal residual disease in solid tumors

**DOI:** 10.1101/2022.04.07.22273446

**Authors:** Dadasaheb Akolkar, Darshana Patil, Sneha Puranik, Swapnil Puranik, Pratiksha Sunil Nandre, Karishma Raosaheb Sabale, Sachin Apurwa, Harshal Bodke, Navin Shrivastava, Vineet Datta, Stefan Schuster, Jinumary John, Ajay Srinivasan, Rajan Datar

**Author notes:** Corresponding author: Dadasaheb Akolkar.

## Abstract

Real time monitoring of disease status is an essential part of cancer management. The low sensitivity and specificity of serum markers and the constraints and risks associated with radiological scans prompt the need for accurate non-invasive means to monitor minimal residual disease (MRD) in solid tumors. In this study we describe MRD evaluation via profiling of patient-specific gene variants in cell free tumor DNA (i.e., ctMRD). We evaluate the feasibility of this approach for real time monitoring of tumor load dynamics in response to anticancer treatments. We prioritized 162 hot spot mutations for designing ctMRD assays based on literature review. These ctMRD assays were evaluated in 436 plasma specimens with a median of 6 (range 3-18) longitudinal evaluations in a cohort of 48 patients with various solid tumors. In patients with partial radiological response (PR), Mutant Allele Fraction (MAF) showed high correlation (84%) with radiological response and tumor volume (cm^3^) compared to conventional CA markers (53%). Total plasma ctDNA level was significantly higher in patients with 2-5 metastatic sites compared with single metastatic site (P = 0.04) and discriminated patients with stable disease (SD) and progressive disease (PD) from patients with partial response (PR) (*P =0.01* and P = 0.04, respectively). Collectively, the present study shows that changes in mutation burden evaluated using patient specific ctMRD assays is a highly sensitive approach for monitoring of therapy response.

## Introduction

Real-time assessment of disease status and treatment response is crucial for cancer management. Early indication of disease progression or recurrence can initiate effective clinical measures to manage the disease. Currently used imaging methods for assessment of treatment response such as magnetic resonance imaging (MRI) ^1^, computed tomography (CT), positron emission tomography-CT (PET-CT) ^2, 3^ or and PET-CT are unsuitable for high frequency use owing to radiation exposure risks and limitations in visualizing micro metastatic disease ^4, 5^. Imaging scans may often be prone to interference from local inflammation, postsurgical changes and necrotic damage ^6^. While serum tumor markers are minimally invasive and less expensive ^7^, they are prone to lower sensitivity and lower specificity in addition to high intra-individual and inter- individual variability ^8, 9^. There is hence a need to develop non-invasive technologies for effective monitoring of MRD and early detection of recurrence.

Circulating Tumor DNA (ctDNA) fragments released following apoptosis of tumor cells is regarded as a sensitive and specific biomarker due to its ubiquity across solid tumors ^10^. Presently, detection of gene features / variants in ctDNA is used to inform selection of targeted anti-cancer agents in cancers such as Non-Small Cell Lung Cancer and Breast Cancer ^11^. For example, BRAF inhibitor Encorafenib for BRAF V600E mutated colon and rectal cancers; tyrosine kinase inhibitors Alectinib, Brigatinib, Lorlatinib, Ceritinib, Crizotinib for ALK rearrangement positive non-small cell lung cancers; NTRK inhibitors Larotrectinib and Entrectinib for NTRK fusion positive solid tumors; anti-EGFR tyrosine kinase inhibitors Erlotinib, Gefitinib, Afatinib, Dacomitinib and Osimertinib for certain EGFR-mutant non-small cell lung cancers. A significant challenge for ctDNA based testing is the reliable detection of gene variants against the large background of normal DNA released by apoptotic non-malignant cells. The sensitivity of detection of rare somatic mutations in the cell free DNA varies with the platform used. For instance, NGS Assay variants such as tagged-amplicon deep sequencing (TAM seq), BEAMing and use of molecular tags can reach detection of Mutant Allele Fraction (MAF) to 0.1% whereas, the digital PCR can reach to 0.01% ^12–14^. However, lack of guidance related to the selective use of these technologies affects their integration into routine patient care for detection on minimal residual disease in solid tumors. While NGS offers multigene testing that encompass tumor heterogeneity and assists in monitoring of tumor clones, digital PCR based ctMRD assays on the other hand offers ultrasensitive detection of the tumor driver clone/s.

The present study focuses on the serial evaluation of patient-specific ctMRD assays and its concordance with radiological status of the disease in a cohort of 48 patients with either of 16 types of malignancies. The predictive value of ctDNA was evaluated based on concordance with radiologically assessed changes to tumor volume. The study shows that changes in ctDNA levels detected via determining specific MAF of targeted loci using ctMRD assays correlates with the treatment responses and supports the role of ctDNA assays for MRD purpose. In addition, we also show that total plasma ctDNA levels were significantly higher in patients with 2-5 metastatic sites as compared to those with single metastatic site; evaluation of plasma cfDNA levels was able to differentiate patients with stable disease (SD) or progressive disease (PD) from patients with partial response (PR).

## Materials and Methods

### Patients

Patients from RESILIENT trial (Trial registration number CTRI/2018/02/011808) (a single arm, single center, non-randomized phase II/III prospective trial for evaluation of treatment response to therapy based on Encyclopedic Tumor Analysis (ETA) recommendation in r/r m-cancers patients) ^15^ were retrospectively analyzed in the present study. The study was approved by the Ethics Committees of both the participating institutes i.e. Datar Cancer Genetics and HCG Manavata Cancer Centre. All patients gave written informed consent. Patients were studied with paired blood collections and scan assessments performed before treatment initiation and at treatment response evaluation.

### Specimen collection

10-15 ml peripheral blood was collected in BD ETDA Vacutainer® (BD USA) from each patient before treatment and at the time when therapy response was assessed. Plasma was extracted within two to three hours of blood collection and was immediately frozen at -80°C till further processing. Fresh resected tumor tissues or biopsies were collected in RNA later^TM^ solution (Ambion) and shipped to laboratory within 2-3 h for ETA analysis.

### NGS workflow

Fresh or FFPE tumor tissue DNA was extracted using The PureLink® Genomic DNA Mini Kit and MagMAX FFPE DNA isolation kit (Thermo Fisher Scientific, USA) as per the manufacturers’ instructions. ctDNA obtained from plasma was used for enrichment of targeted regions of 52 genes using Ion Oncomine Pan Cancer Panel and tumor DNA for 452 genes using Oncomine Comprehensive Assay v3 and Ion AmpliSeq Comprehensive Cancer Panel (Thermo Fisher Scientific, USA) as per user recommended protocols. One hundred micromoles of library were further subjected to template preparation with Ion OneTouch 2 and sequenced on Ion PI semiconductor chip (Thermo Fisher Scientific, USA). Primary data processing was carried out using the Torrent Suite Software (version 5.10) on a Torrent Server. Variant calling was performed using the Ion Reporter Software (version 5.10) (Thermo Fisher Scientific, USA).

### Design and optimization of patient specific assays

The Single Nucleotide Alterations (SNA) detected by the genomic analysis were considered for designing patient-specific ctMRD assay using Custom TaqMan® Assay Design Tool (Thermo Fisher Scientific, USA) (Supplementary Table S1). Plasmids harboring mutant sequences were custom designed using GeneArt Gene synthesis service (Invitrogen, CA, USA). Assay optimization was performed for the ctMRD assays which gave aberrant mutant positive droplets in wild type only controls at recommended annealing temperature (Supplementary Table S2). Input concentration of 50 ng of wildtype DNA (Control DNA CEPH 1347-02, Thermo Fisher Scientific, USA) was used per reaction (this was the maximum amount of DNA input to be used for an assay) and the temperature was varied across the wells by generating a thermal gradient PCR T100 (Bio-Rad Laboratories, USA). The temperature which provided largest fluorescent amplitude separation of wild type copies without any mutant positive droplets, was considered optimal for each ctMRD assay.

ctDNA was extracted from 2-4 ml plasma using QIAamp Circulating Nucleic Acid Kits (Qiagen, Germany). ctDNA was quantified by Qubit dsDNA HS Assay Kit with the Qubit Fluorometer (Thermo Fisher Scientific, USA). ctDNA was additionally quantified by means of droplet digital PCR (ddPCR) system from Bio-Rad QX200 (BIORAD, USA). Dual labeled (FAM or HEX) fluorescent probes were used for the mutant and wild type loci PCR, respectively. The droplet generation was carried out in QX200 AutoDG Droplet Digital PCR System according to manufacturer’s instructions (Instruction Manual, QX200 AutoDG Droplet Digital PCR System –Bio-Rad). The reaction mixtures were subsequently analyzed using the QX200 reader, according to manufacturer’s instructions. The mutant allele fraction (MAF), the fraction of mutant droplets among the total number of mutant and wild-type droplets was estimated using QuantaSoft v.1.7.4 software (Bio-Rad, USA). Patient specific mutant alleles were selected based on higher MAF and the presence of concordance of mutant allele between ctDNA and tissue (Supplementary Table S1).

### Tumor burden and response assessment

Tumor burden estimation was performed with computed tomography (CT), or positron-emission tomography coupled with CT (PET-CT). Scans were evaluated by a radiologist as per Response Evaluation Criteria in Solid Tumors version 1.1 (RECIST 1.1) ^16^. Tumor volume measurements were also performed assuming ellipsoid shape of the tumor and mentioned as Centimeter^3^ (cm^3^) as described previously ^17^. The imaging measurement of tumor response was qualitatively classified into four categories: complete response (CR), partial response (PR), stable disease (SD) and progressive disease (PD). The maximum interval between blood biomarker analysis and radiological assessment was 4 weeks for baseline and treatment response follow-ups.

### Measurement of Tumor Markers

Serum CEA, CA19-9, CA 15-3, LDH and CA125 measurements were performed on plasma specimens from Thyrocare Laboratories, Mumbai. All CA markers were tested prior to initiation of chemotherapy (within 7 days) and every 4 weeks after initiation of chemotherapy depending on availability of serum specimens for respective time points. The CA marker data for all patients evaluated and cut-off values for all the markers are described in Supplementary Table S3.

### Statistical analysis

Pearson test was used to assess a pairwise correlation between MAF, CA marker, ctDNA concentration and tumor burden during treatment response reassessment. P-values ≤ 0.05 were statistically significant. Statistical analyses were performed using GraphPad Prism 7.0 (GraphPad Software Inc., San Diego, CA, USA).

## Results

### Somatic multigene profiling of tumor tissues and ctDNA

Total 78 patients with tumor and plasma specimens were included in the study with different cancer types: HNSCC (n=19, 24%), breast cancer (n=16, 20%), colorectal (n=8, 10%), lung cancer (n=7, 9%), cervix and pancreatic (n=5, 6.4% each), ovarian cancer (n=4, 5%), gastric and hepatobiliary (n=3, 4% each), prostate (n=2, 2.6%), esophageal, duodenum, yolk sac tumor, testicular cancer, synovial sarcoma and medulloblastoma (n=1, 1.3% each) (Table 1, Figure 1).

**Figure 1.**
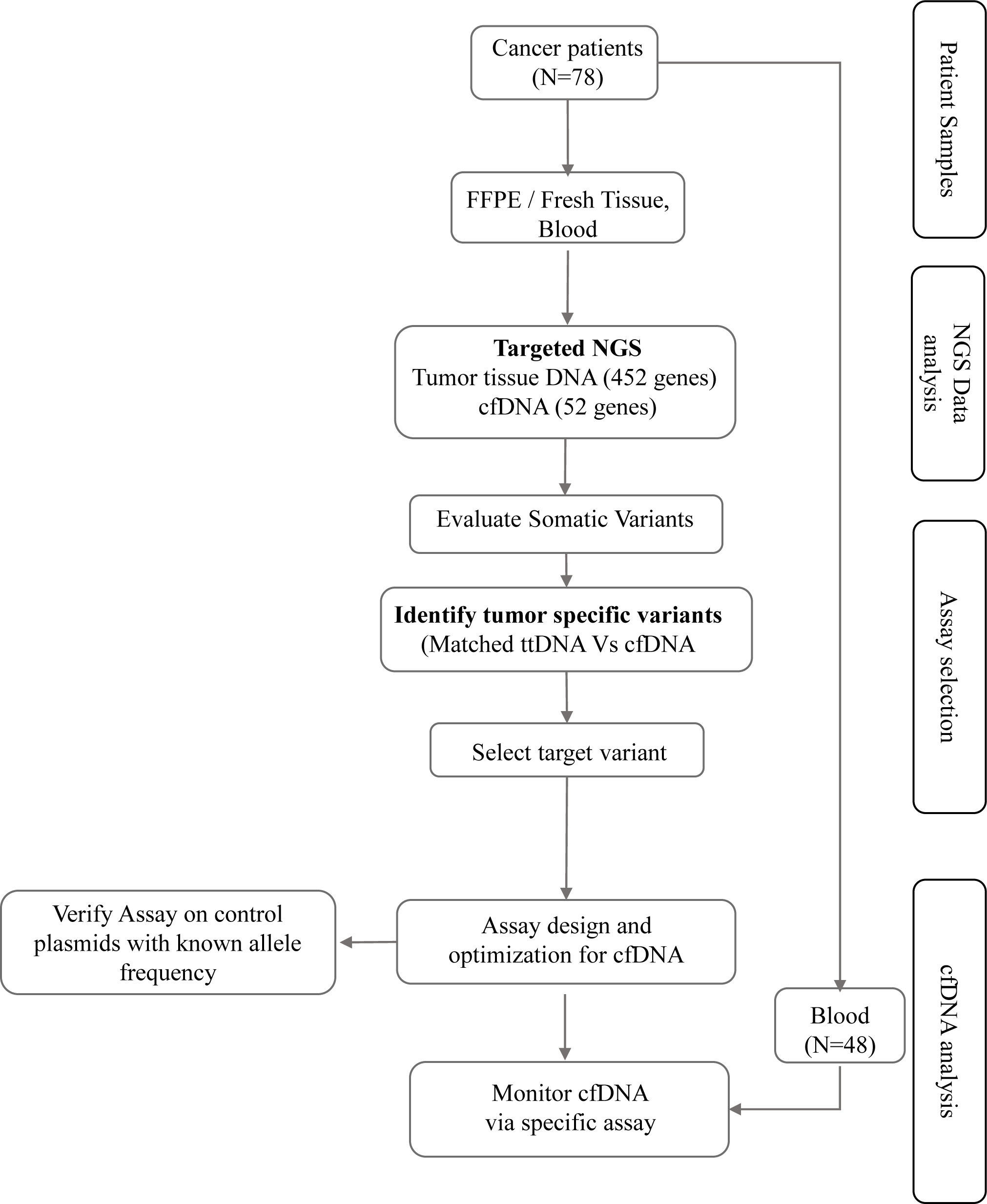
Workflow of the study.

**Table 1.** Patients’ Characteristics

| **Table 1: Clinical characteristics (N=78)** |  |
| --- | --- |
| **Characteristics** | **N (%)** |
| **Age** |  |
| Median years (range) | 50 (8 - 79) |
| **Gender** |  |
| Female | 40 (51) |
| Male | 38 (49) |
| **Cancer type/organ** |  |
| HNSCC | 19 (24) |
| Breast | 16 (20) |
| CRC | 8 (10) |
| Lung | 7 (9) |
| Cervix | 5 (6.4) |
| Pancreas | 5 (6.4) |
| Ovary | 4 (5) |
| Gastric | 3 (4) |
| Hepatobiliary | 3 (4) |
| Prostate | 2 (2.6) |
| Esophageal | 1 (1.3) |
| Duodenum | 1 (1.3) |
| Medulloblastoma | 1 (1.3) |
| Yolk sac tumor | 1 (1.3) |
| Synovial Sarcoma | 1 (1.3) |
| Testes | 1 (1.3) |
| **Disease status** |  |
| **Grade** |  |
| I | 5 (6.4) |
| II | 25 (32) |
| III | 37 (47.5) |
| IV | 1 (1.3) |
| Unknown | 10 (13) |
| **Stage** |  |
| IV | 78 (100) |
| **Metastasis (Number of sites)** |  |
| 0 | 2 (2.6) |
| 1 | 29 (37) |
| 2 | 25 (32) |
| 3 | 13 (17) |
| 4 | 5 (6.4) |
| 5 | 2 (2.6) |
| Unknown | 2 (2.6) |
| **Prior treatment** |  |
| Median Number of lines (range) | 4 (1 - 17) |
| Abbreviation: HNSCC = Head and neck squamous cell carcinoma, CRC = colorectal cancer |  |

Genomic profiling of tumor specimens by deep NGS sequencing in 78 patients revealed median of 4 (range 1-25) alterations based upon analysis of 452 genes (Supplementary Table S4) in the targeted region of 1.8 Mb (Figure 2). TP53 was found to be the most commonly mutated in 59 cases (76%) followed by PIK3CA in 29 (37%), EGFR in 22 (27%), GNAS and KRAS in 18 each (23%) and PDE4DIP in 15 each (19%), respectively. Mutations in other cancer associated genes were identified in the range of 2 to 20% (Figure 2). Paired mutation analysis of plasma derived ctDNA through analysis of 52 genes (Supplementary Table S5) using molecular tag-based sequencing, identified non-synonymous somatic mutations. The mutation profiles for patients harboring ctDNA mutations are shown in Figure 2 (yellow rectangles with black circles).

**Figure 2.**
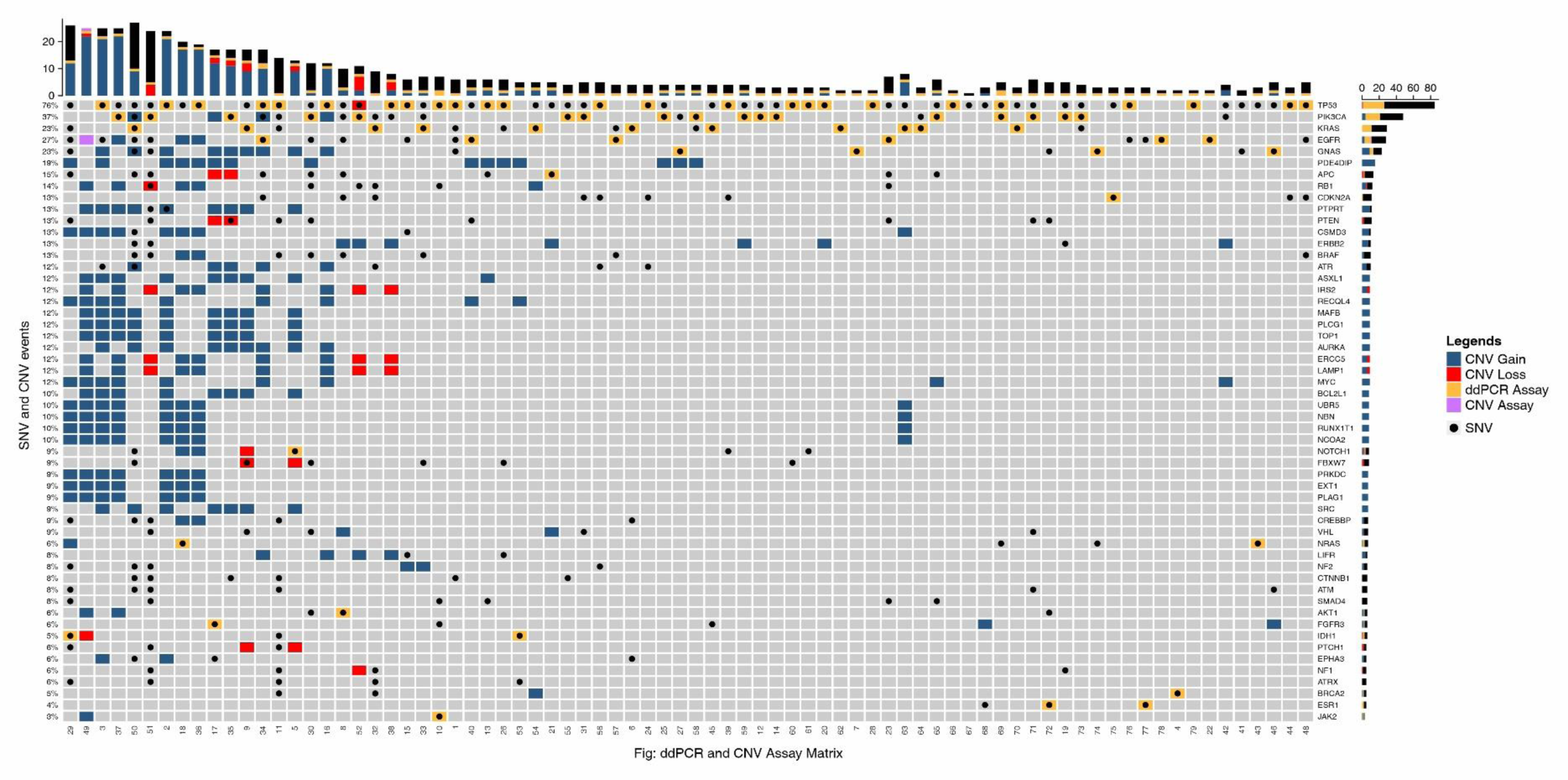
Molecular profiles for 78 cancer patients showing single nucleotide variations (SNVs) and copy number variations (CNVs). Black circle indicates SNV analysis by Next Generation Sequencing (NGS), Blue and red boxes are for CNV gain and loss, respectively. Black circles overlapped with yellow boxes represent ddPCR assays designed for respective SNVs detected in ctDNA.

### Selection of the ctMRD Assays

Forty eight patients were selected to monitor the treatment outcome and the criteria used for selection of ctMRD was based on i) mutation concordance between tumor tissue and ctDNA (n=18/48 patients; 37.5%), ii) high MAF followed by presence of mutation exclusively either in ctDNA (n=17; 35.5%) or iii) tumor tissue (n=13; 27%) (Supplementary Table S1). The selected assays were optimized for better separation of wild type and mutant droplets as well as false positive rate of respective assay was determined (Figure 1 workflow, Supplementary Figure S1). For instance, assays which gave aberrant mutant positive droplets in wild-type DNA controls (no mutant DNA spiked) at recommended annealing temperatures were optimized by varying annealing temperatures. JAK2 V617F assay (recommended annealing temperature 60° C) and IDH1 R132H assay (recommended annealing temperature 55°C) were optimized at 60.8°C and 56.5°C, respectively (Supplementary Figure S1A-B). The limit of detection was determined for respective assays by spiking mutant and wild-type alleles to obtain MAF of 0.005%, 0.01%, 0.05% and 0.5% (Supplementary Figure S1C). Thus, the optimized assays (Table S2) were employed for serially monitoring patient specific mutation from ctDNA at 0.01% LOD to evaluate treatment outcome. Serial monitoring was performed using selected ctMRD assay on median number of 6 follow ups (range 3 to 18) (Figure 2).

### Monitoring tumor burden following therapy via ctMRD assay

Forty patients out of 48 with different types of solid malignancies (Table 2) had received baseline and reassessment PET scans along with CA markers and ctDNA evaluations were analyzed for tumor response. In a cohort of breast cancer patients (n=7), longitudinal blood specimens collected after one course of chemotherapy were analyzed for respective ctMRD assay in plasma derived ctDNA (Supplementary Table S1). Among these, 3 patients had mutation in PIK3CA gene, 2 were with TP53 mutation and two with GNAS mutation. Clinical response was recorded as PR in 3 patients (12, 14, and 3) which showed decreased tumor volume with concomitant decrease in MAF (Figure 3A-C). Patient 14 showed PR till ∼5 months followed by increased trend of tumor volume as well as MAF from ∼6 months onwards implicating disease progression (Figure 3B). Stable disease evaluated in remaining four patients (SD as patient never achieved below -30% change in SLD) showed consistent trend in MAF with tumor burden (Figure 3D-G).

**Figure 3.**
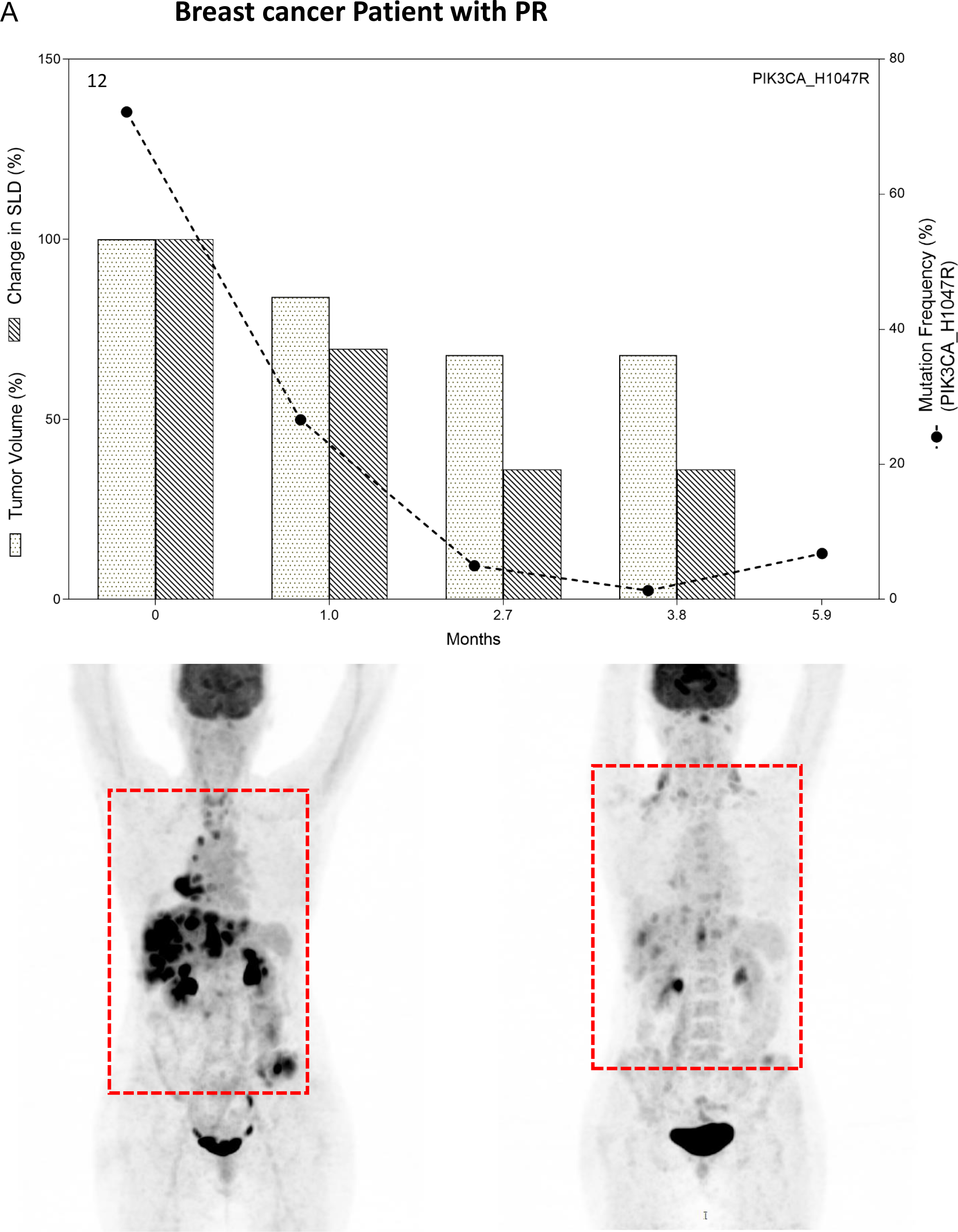
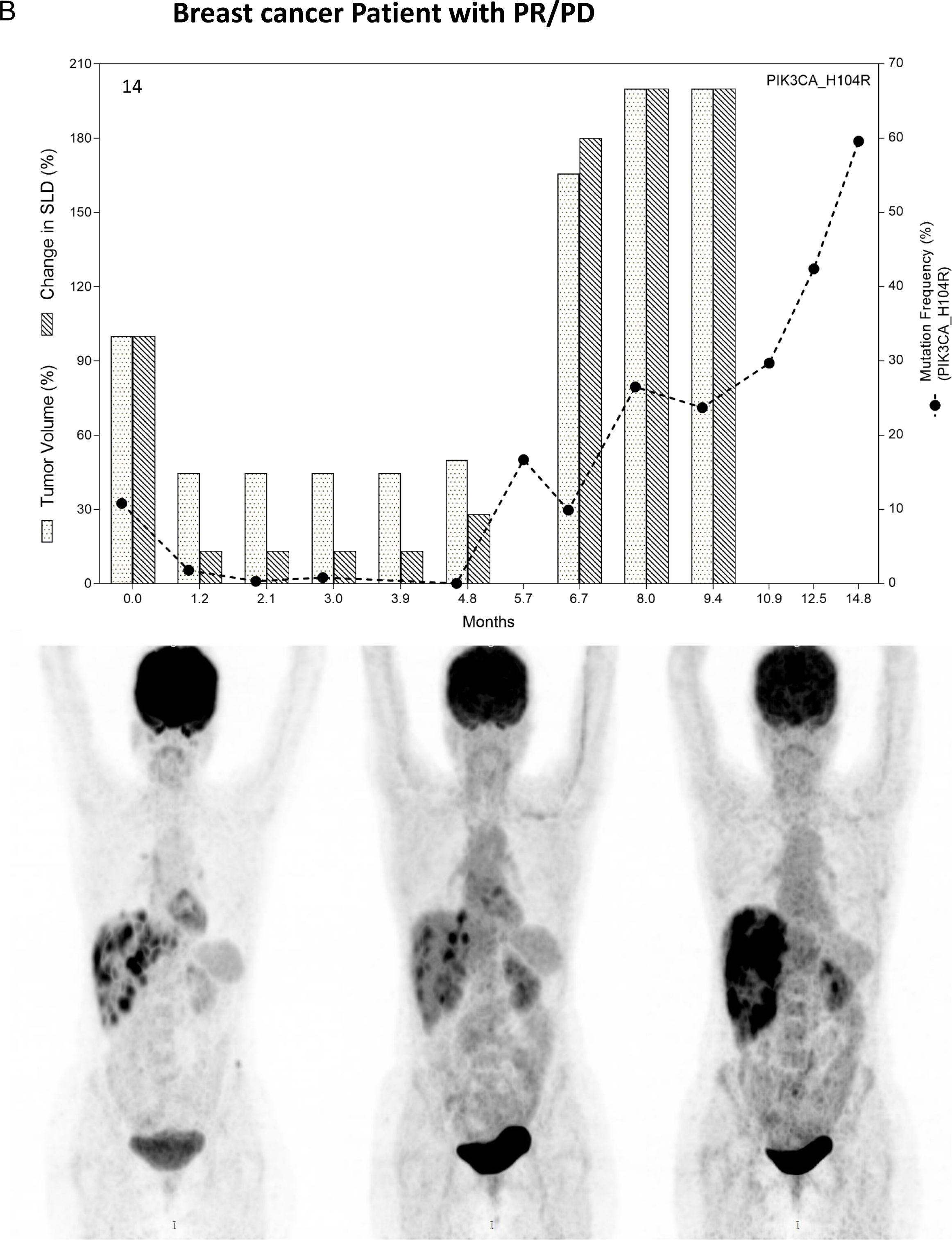
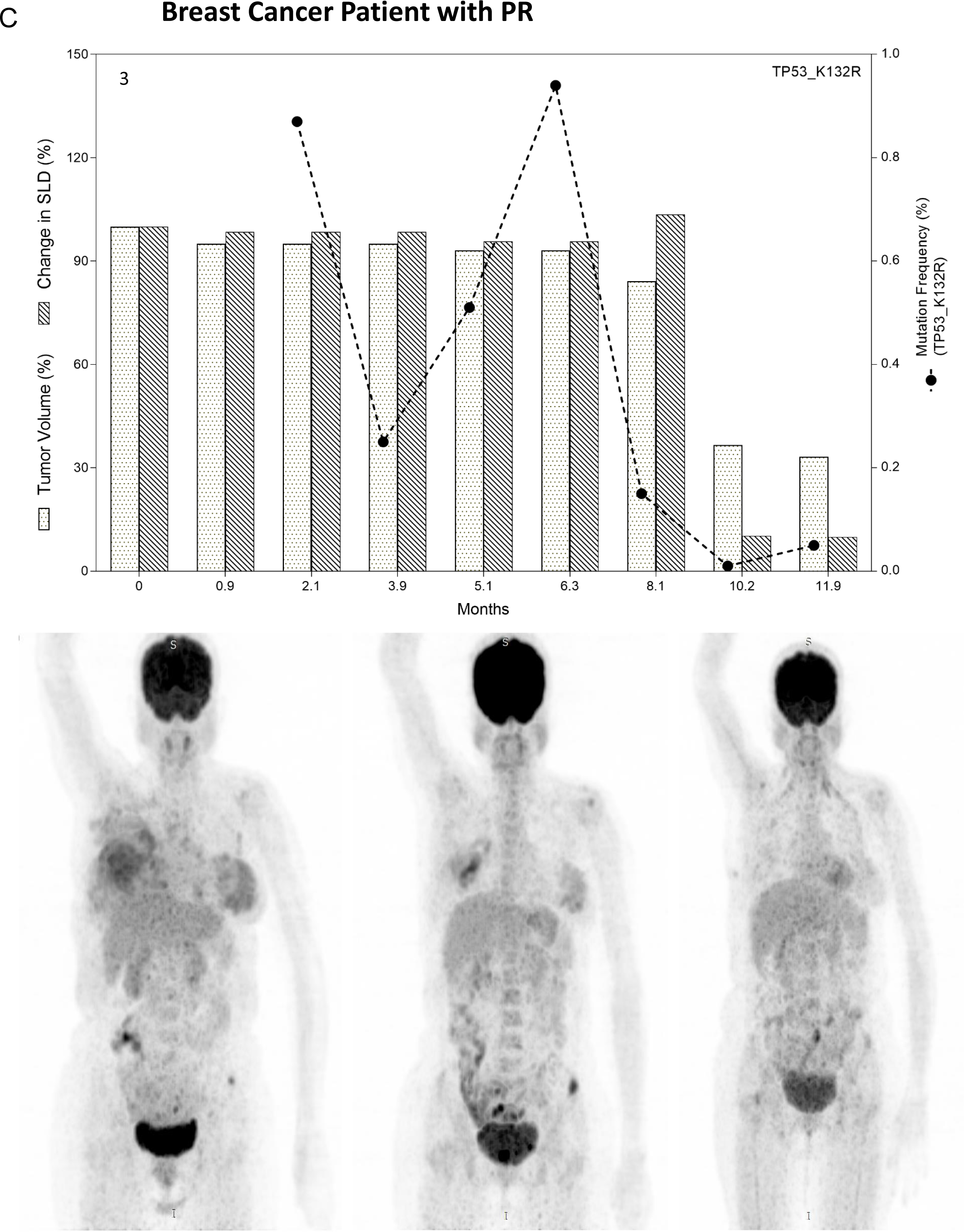
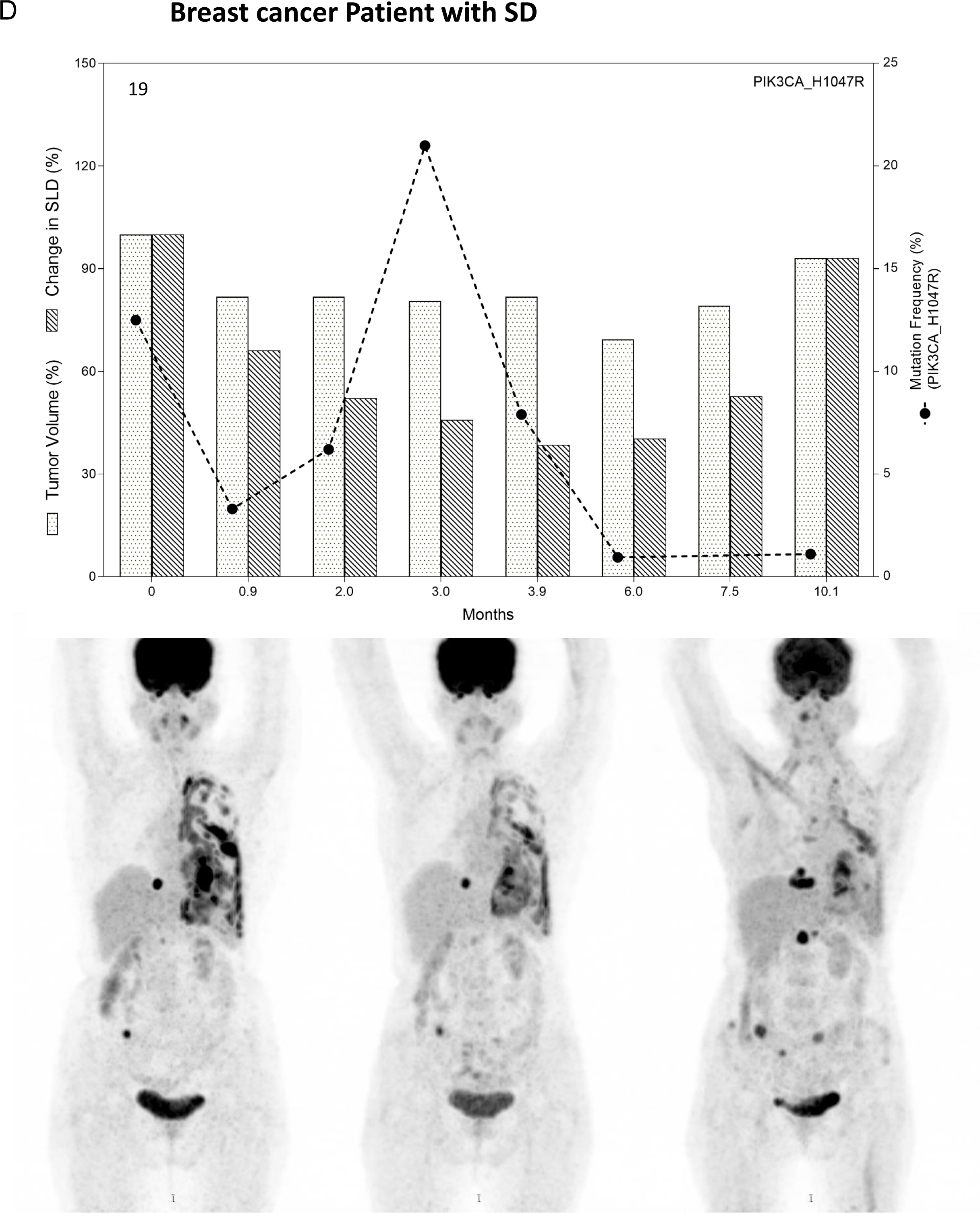
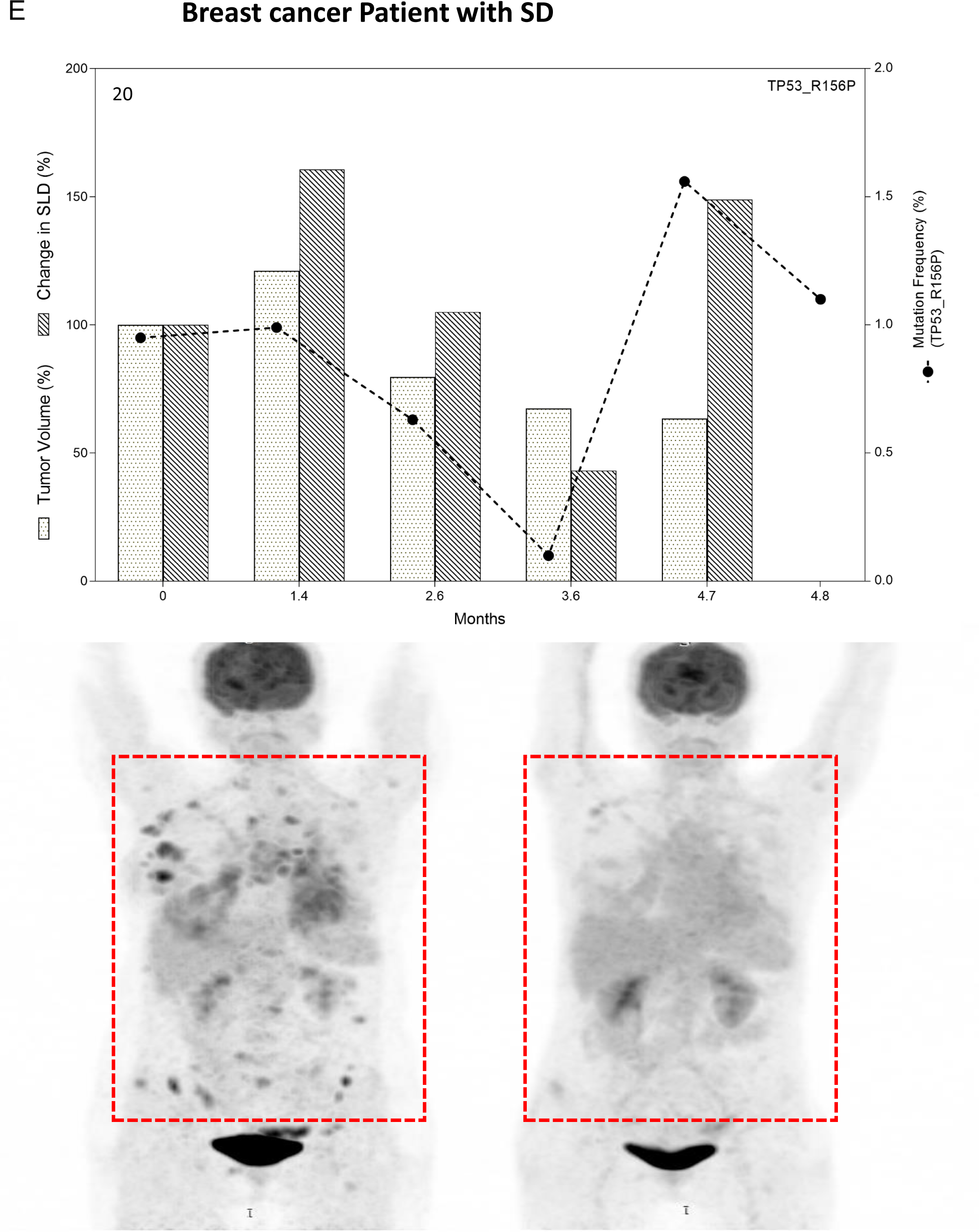
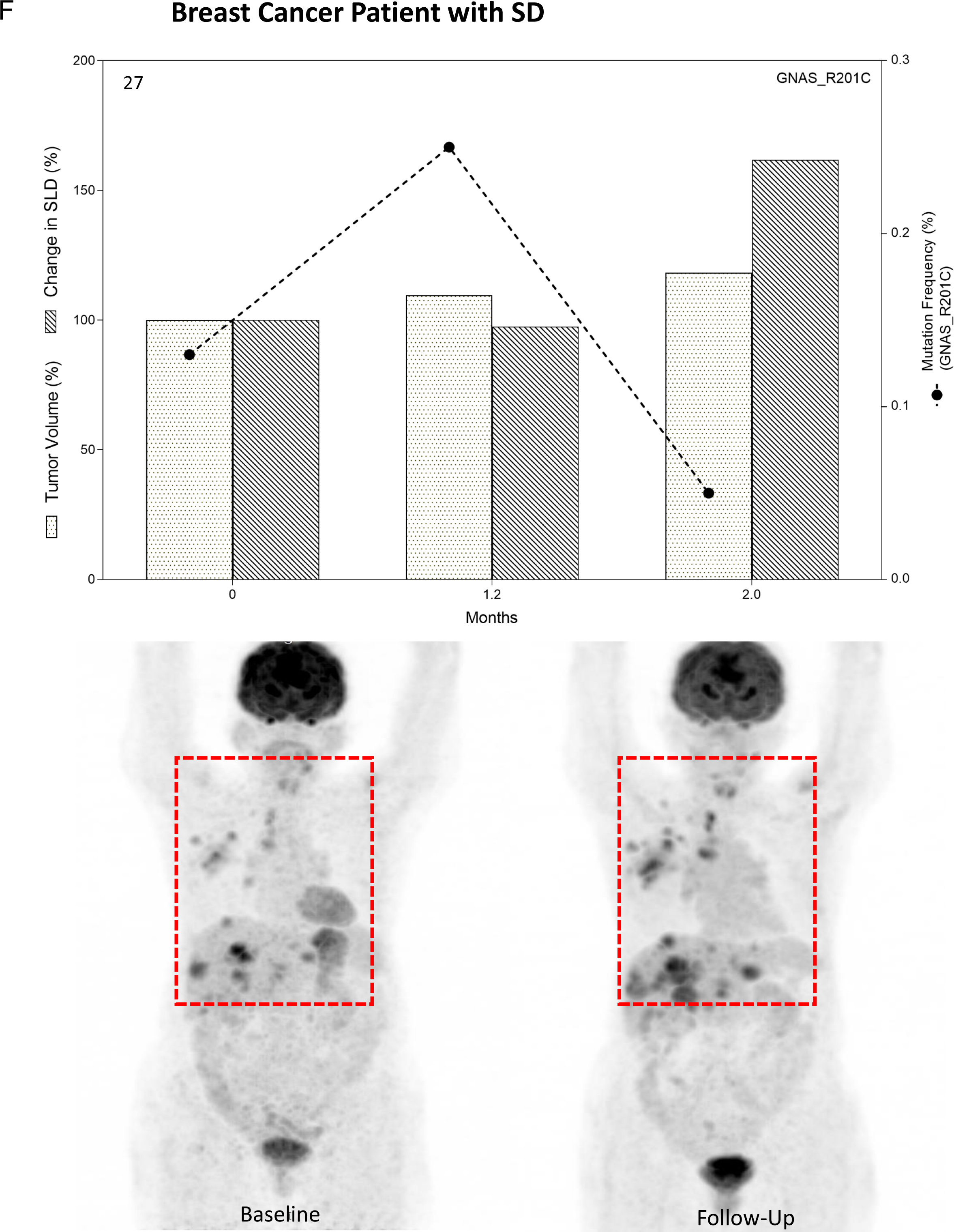
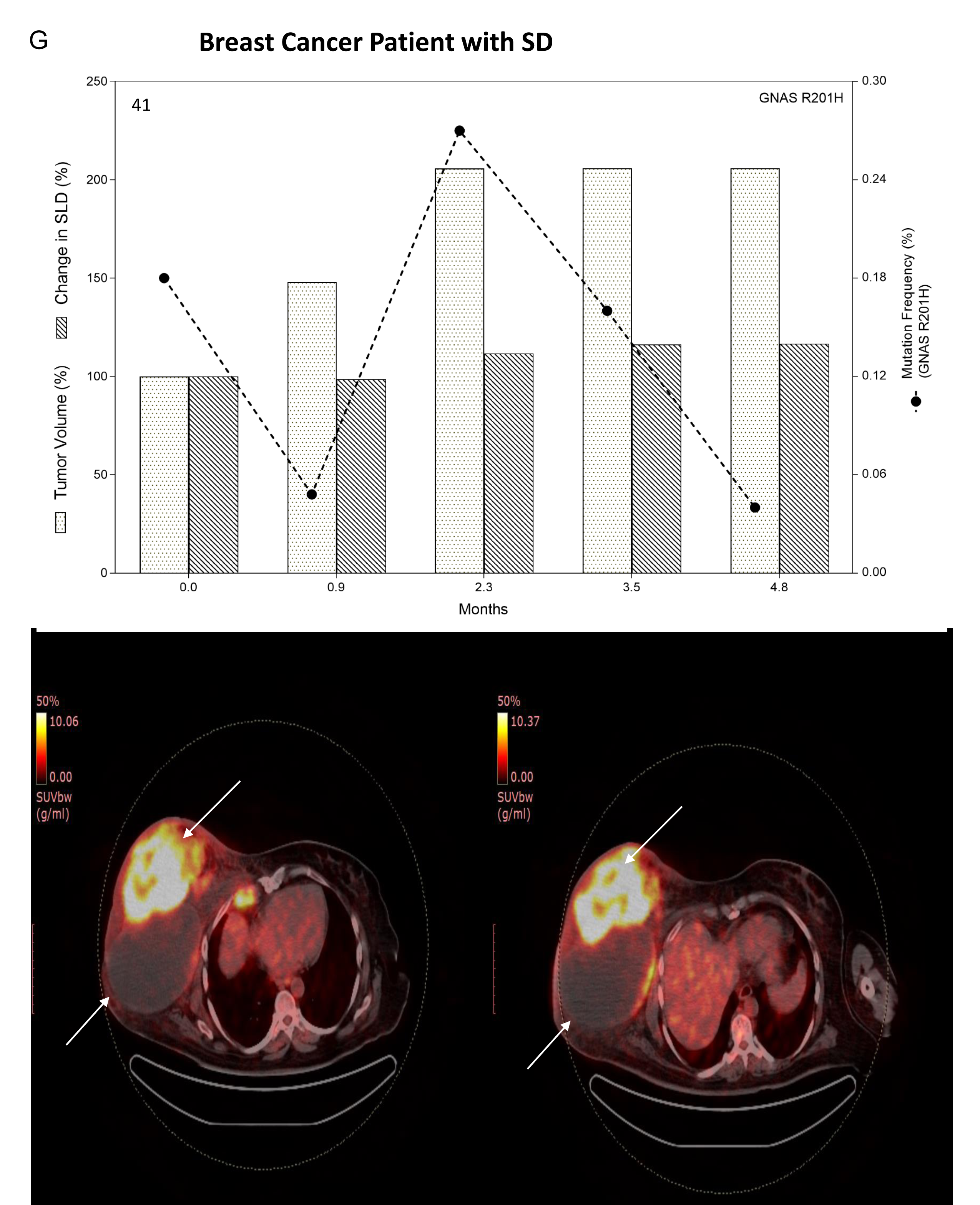
Monitoring Tumor Response by analyzing patient specific hot spot mutations by ddPCR in Breast cancer patients (A - G). Representative coronal views / transverse sections from PET-CT images of patients at Baseline and Follow-Up scan are shown in lower panel of respective graph. Tumor measurements were by RECIST 1.1 criteria (Single lesion dimension (SLD %) and ellipsoid tumor volume (cm^3^) showed on Y-axis. Respective MAF (Mutant allele fraction) from ctMRD assay are on Z-axis with the follow-up time in Months on X-axis.

**Table 2.** Pearson’s correlation analysis between MAF, CA marker, ctDNA concentration and tumor volume (P-values are mentioned in brackets).

| **Table 2: Correlation analysis with tumor volume represented as Pearson's correlation coefficient 'r' (P-value)** |  |  |  |  |  |
| --- | --- | --- | --- | --- | --- |
| **Cancer Type/organ** | **Patient no.** | **MAF** | **CA marker** | **cfDNA conc.** | **Disease outcome** |
| Breast | 3 | **0.6 (0.04)** | **0.9 (0.004)** | -- | PR |
|  | 14 | **0.92 (0.04)** | **0.85 (0.02)** | **0.96 (0.003)** | PR/PD |
|  | 20 | 0.85 (0.09) | **0.2 (0.001)** | -- | SD |
|  | 19 | -- | **0.8 (<0.0001)** | **0.6 (0.001)** | SD |
|  | 27 | -- | 0.99 (0.1) | 0.98 (0.1) | SD |
|  | 12 | 0.98 (0.2) | **0.99 (0.05)** | 0.3 (0.1) | PR |
|  | 41 | -- | **0.8 (0.02)** | **0.1 (0.02)** | SD |
| HNSCC | 24 | **0.2 (0.01)** | -- | **0.26 (0.003)** | SD |
|  | 36 | 0.97 (0.5) | -- | -- | PR |
|  | 39 | -- | **1 (0.01)** | -- | SD |
|  | 34 | 0.99 (0.06) | **0.4 (0.01)** | -- | SD |
|  | 48 | **0.6 (0.02)** | -- | -- | PR |
|  | 43 | 0.98 (0.07) | -- | **0.2 (0.008)** | PR |
|  | 44 | **0.98 (0.04)** | **0.7 (0.05)** | -- | PR |
| CRC | 21 | **0.65 (0.01)** | -- | -- | SD |
|  | 18 | -- | **0.2 (0.02)** | -- | PR |
|  | 10 | -- | -- | 0.7 (0.07) | SD |
|  | 9 | -- | -- | -- | SD |
|  | 6 | 0.4 (0.2) | NA | -- | PD |
|  | 38 | 0.87 (0.07) | **0.54 (0.001)** | -- | SD |
| Pancreas | 29 | **0.44 (0.01)** | -- | **0.96 (0.02)** | SD |
|  | 22 | 0.6 (0.06) | -- | **0.8 (<0.0001)** | PR |
|  | 33 | **0.93 (0.02)** | -- | **0.4 (0.05)** | PR |
|  | 45 | 0.3 (0.07) | -- | 0.98 (0.2) | SD |
| hepatobiliary | 25 | **0.4 (0.05)** | **0.7 (0.01)** | -- | PR |
| Cervix | 5 | 0.95 (0.2) | data NA | -- | PR |
|  | 26 | -- | -- | **0.53 (0.01)** | PR |
|  | 13 | **0.9 (0.01)** | -- | -- | SD |
| Ovary | 16 | **0.65 (0.04)** | 0.72 (0.06) | **0.52 (0.007)** | PR |
|  | 4 | -- | 0.88 (0.08) | **0.37 (0.04)** | PD |
|  | 1 | 0.96 (0.3) | **0.84 (0.0006)** | 0.7 (0.1) | PR |
|  | 2 | 0.75 (0.07) | -- | -- | SD |
| Esophageal | 28 | -- | -- | -- | SD |
| Gastric | 15 | -- | -- | -- | SD |
|  | 46 | 0.3 (0.2) | -- | -- | PR |
| Lung | 7 | -- | 0.99 (0.08) | 0.63 (0.06) | SD/PD |
|  | 23 | -- | **0.75 (0.007)** | -- | PR |
|  | 40 | 1 (0.1) | -- | -- | SD |
| Testes | 32 | 0.96 (0.6) | -- | -- | PR |
| Prostate | 42 | **0.72 (0.007)** | 0.33 (0.09) | 0.3 (0.08) | PR |
| Total Patients | (N=40) | (N=27) | (N=19) | (N=18) |  |
| Abbreviation: MAF = Mutant allele fraction, cfDNA = Circulating free DNA, HNSCC = Head and neck squamous cell carcinoma, CRC = colorectal cancer, MAF = mutant allele frequency, NA = data not available, , '--' = No correlation; PR = Partial response; SD = Stable disease; PD = Progressive disease. |  |  |  |  |  |

Furthermore, 7 HNSCC patients were monitored for their treatment response based on TP53 (n=6) and NRAS (n=1) alterations in the ctDNA. Four patients were evaluated with PR and 3 were with SD. Patient 48 with Buccal mucosa cancer showed PR till 6 months followed by trend of disease progression (Figure 4A) while the other patient 36 with same cancer type indicated PR with decreased trend in MAF (TP53 R248Q) (Figure 4B). Patients 43 and 44 with tonsil and alveolus cancer, respectively showed decreased trend in MAF with tumor volume implicating PR (Figure 4C-D). Remaining 3 patients were with SD (Figure 4E-G). Esophageal cancer (patient 28) and gastric cancer (patient 15) cases assessed by TP53 MAF showed SD, with consistent trend in MAF (Figure 5A-B). The second gastric cancer case (patient 46) monitored with GNAS R201C mutation showed PR with significant decrease in MAF at 3.4 months follow-up (Figure 5C).

**Figure 4.**
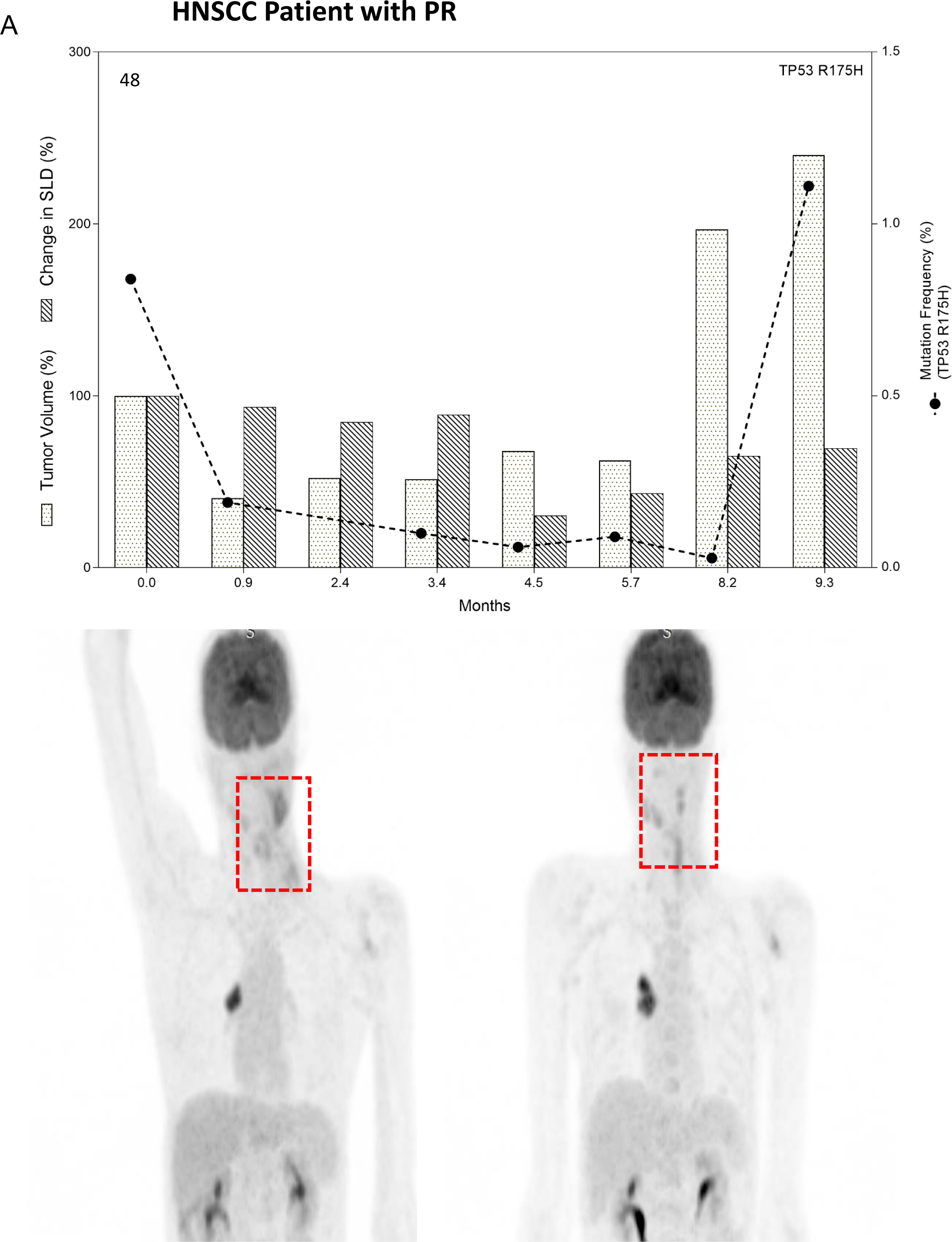
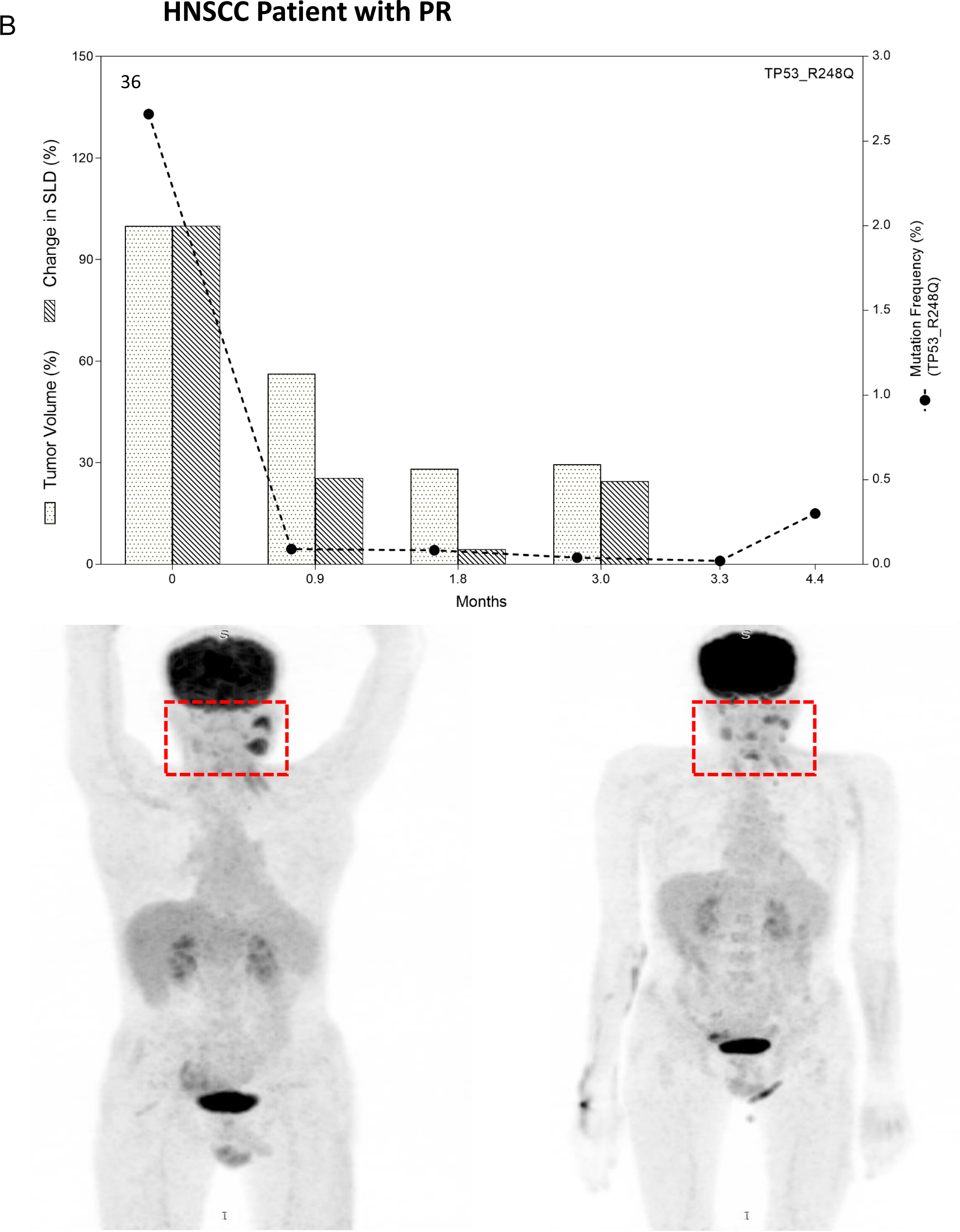
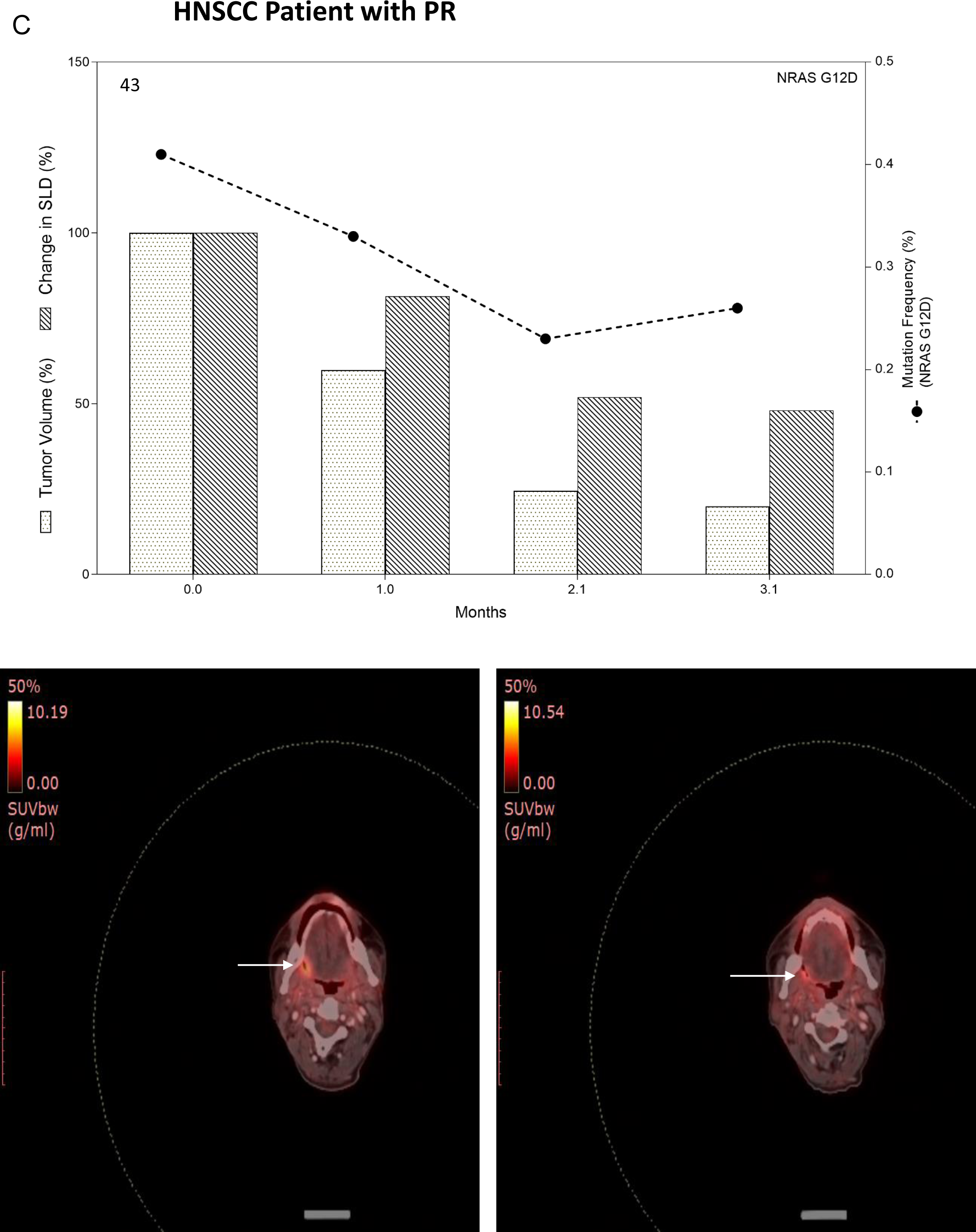
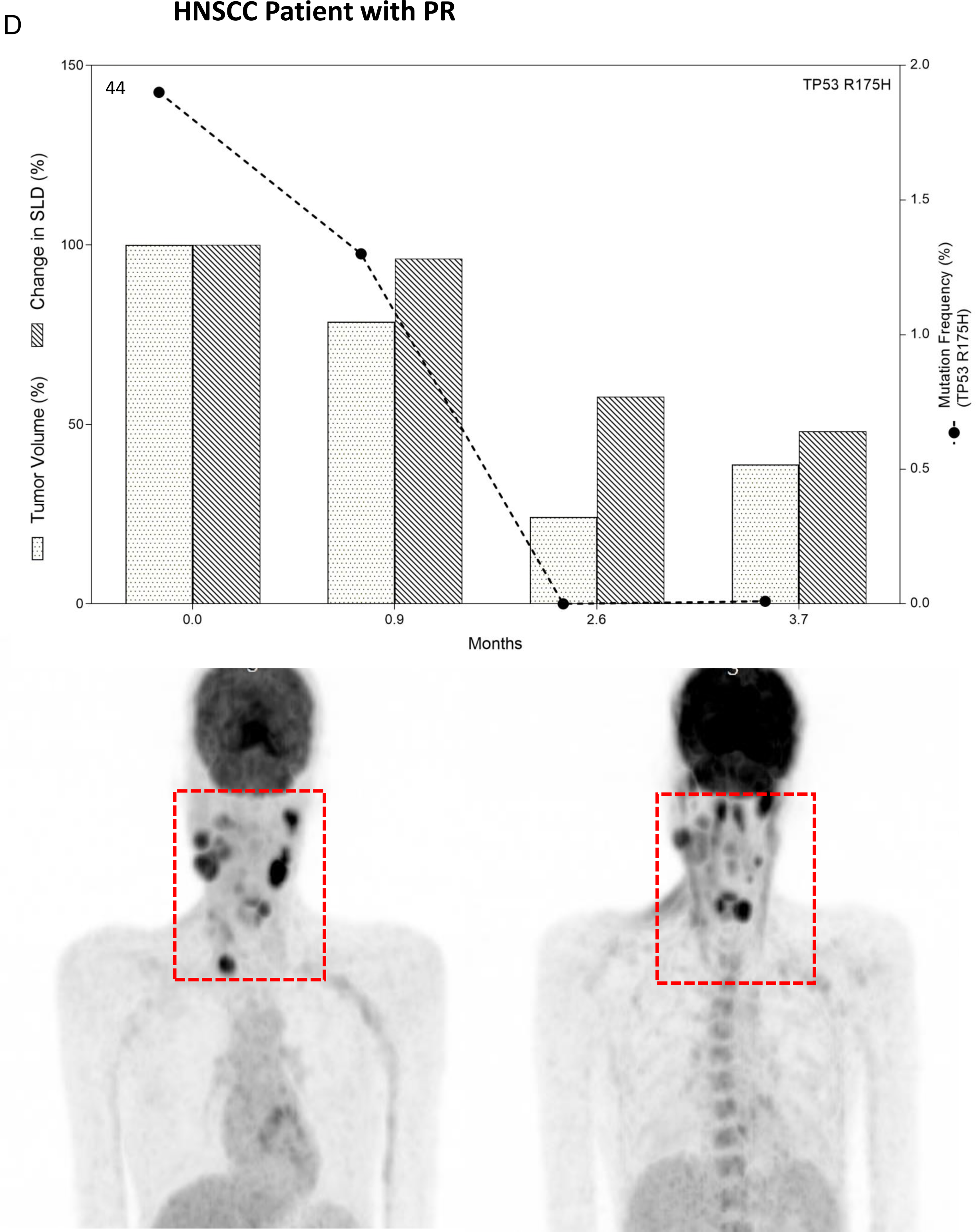
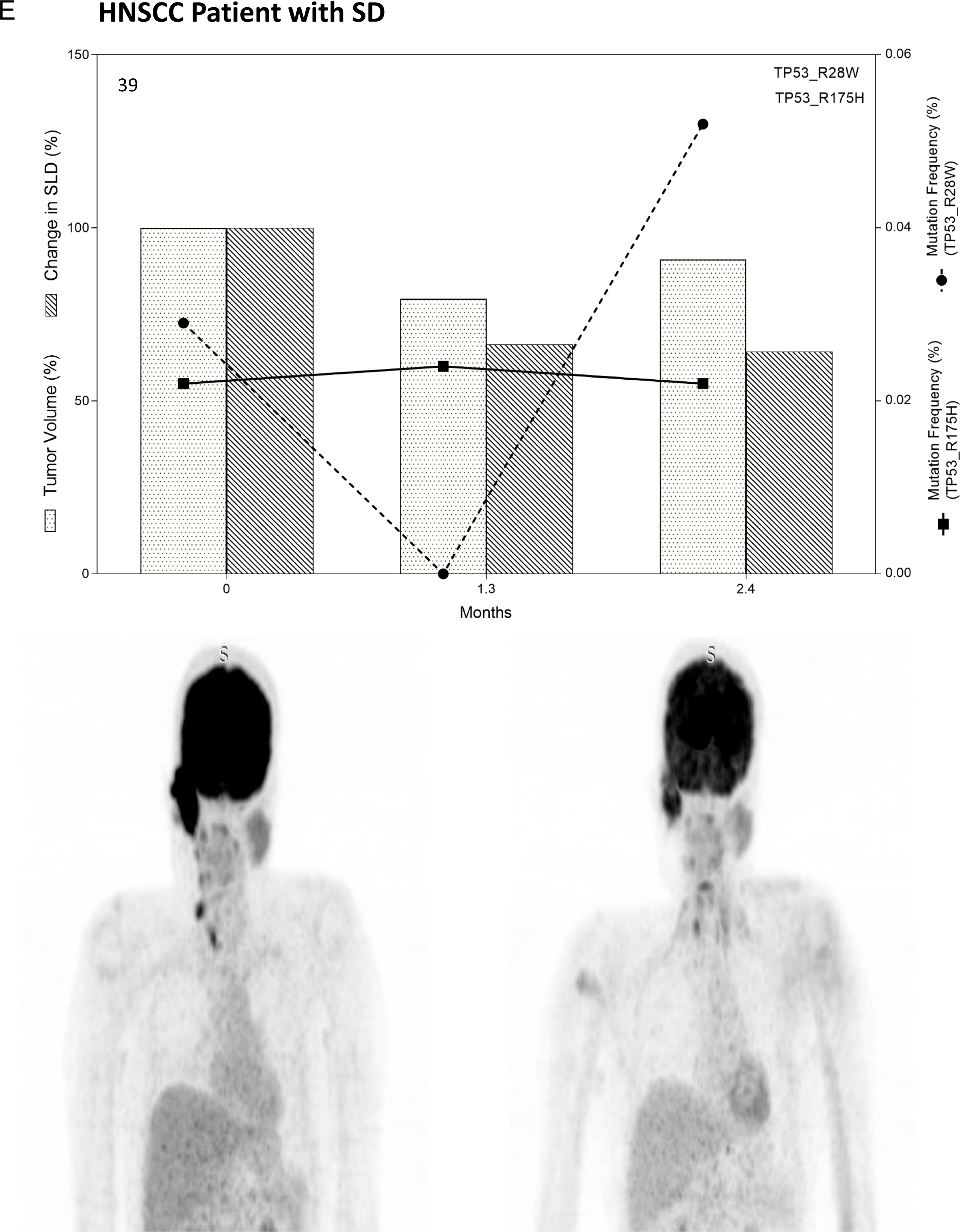
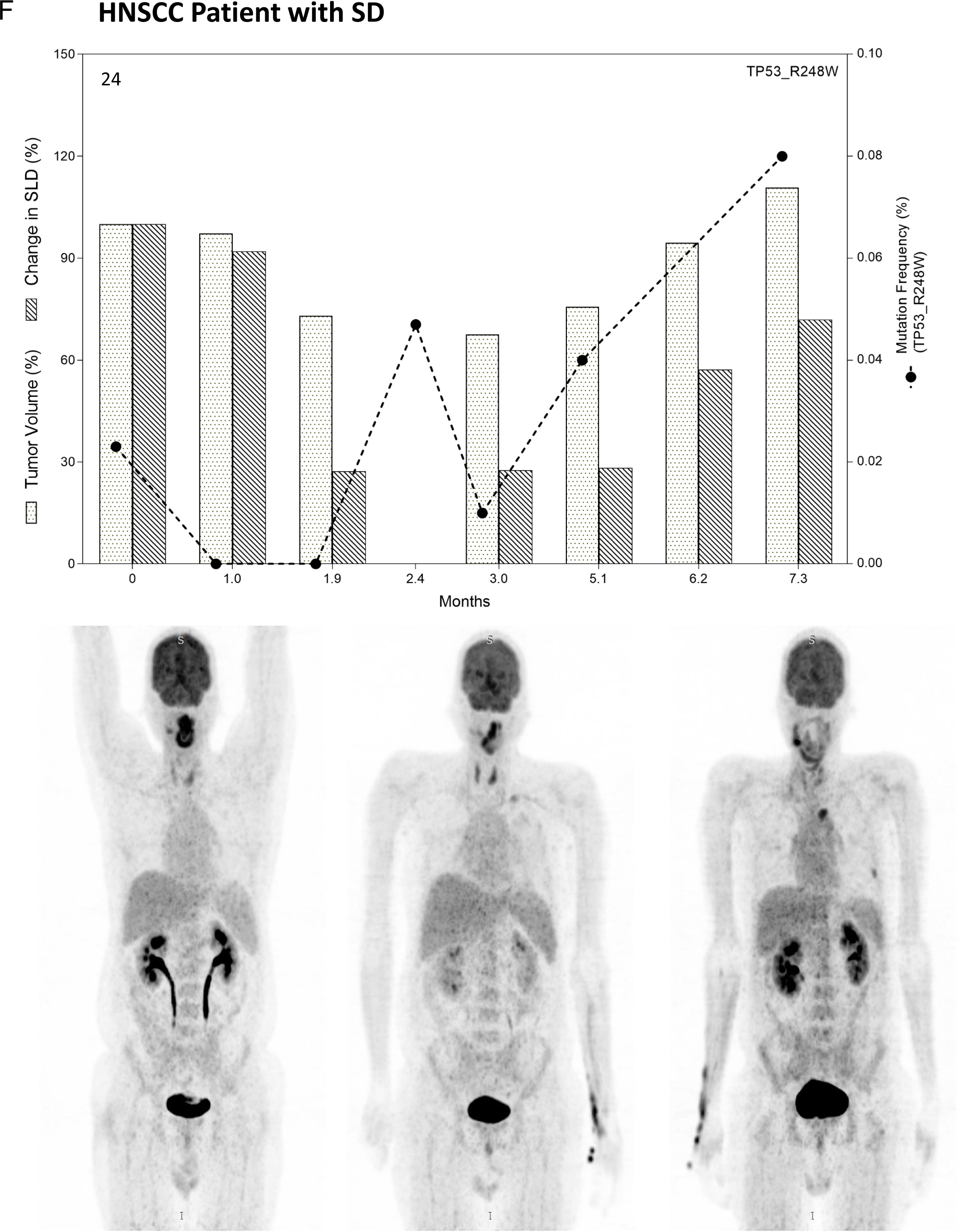
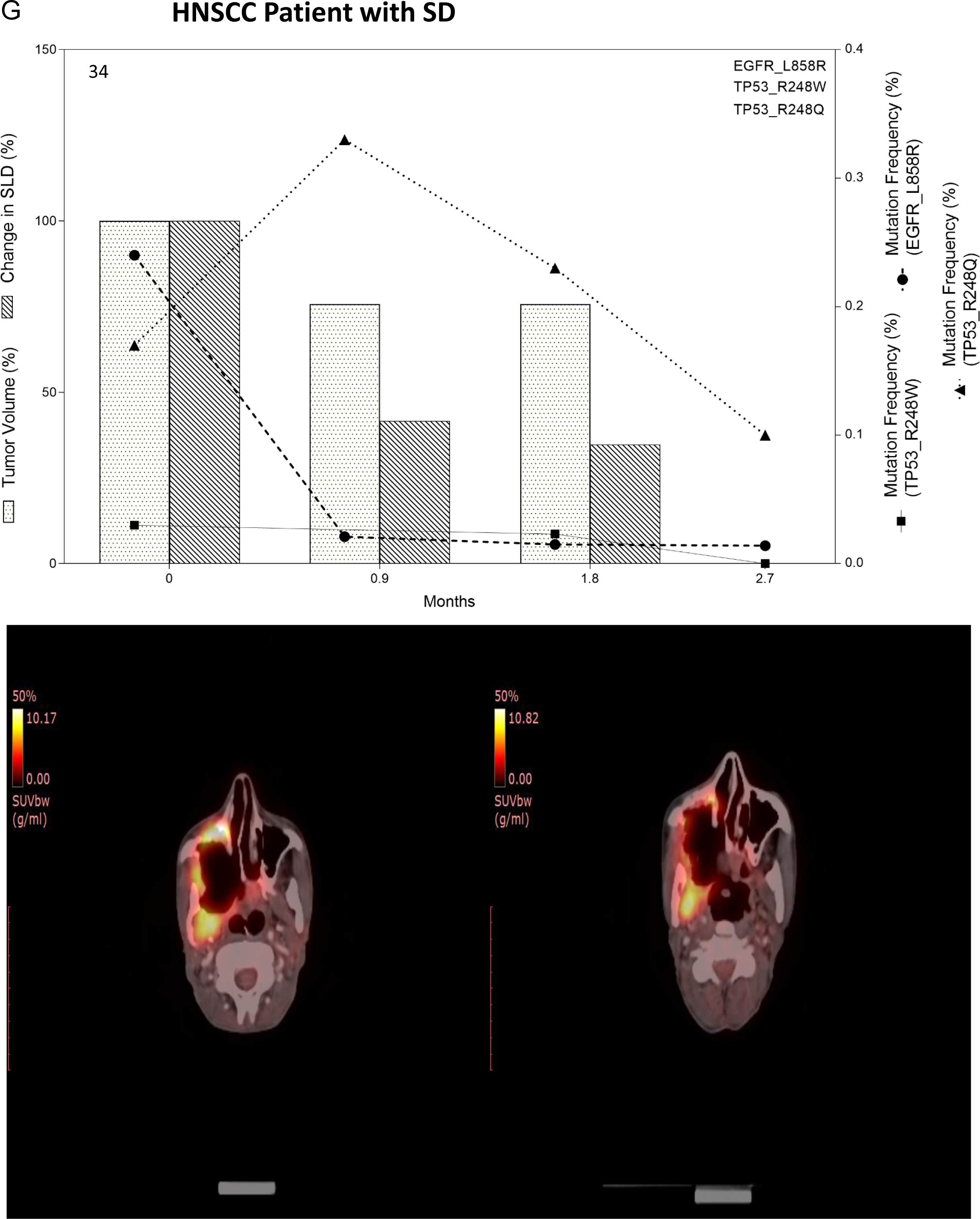
Monitoring Tumor Response by analyzing patient specific hot spot mutations by ddPCR in HNSCC patients (A-G) as mentioned in figure 3.

**Figure 5.**
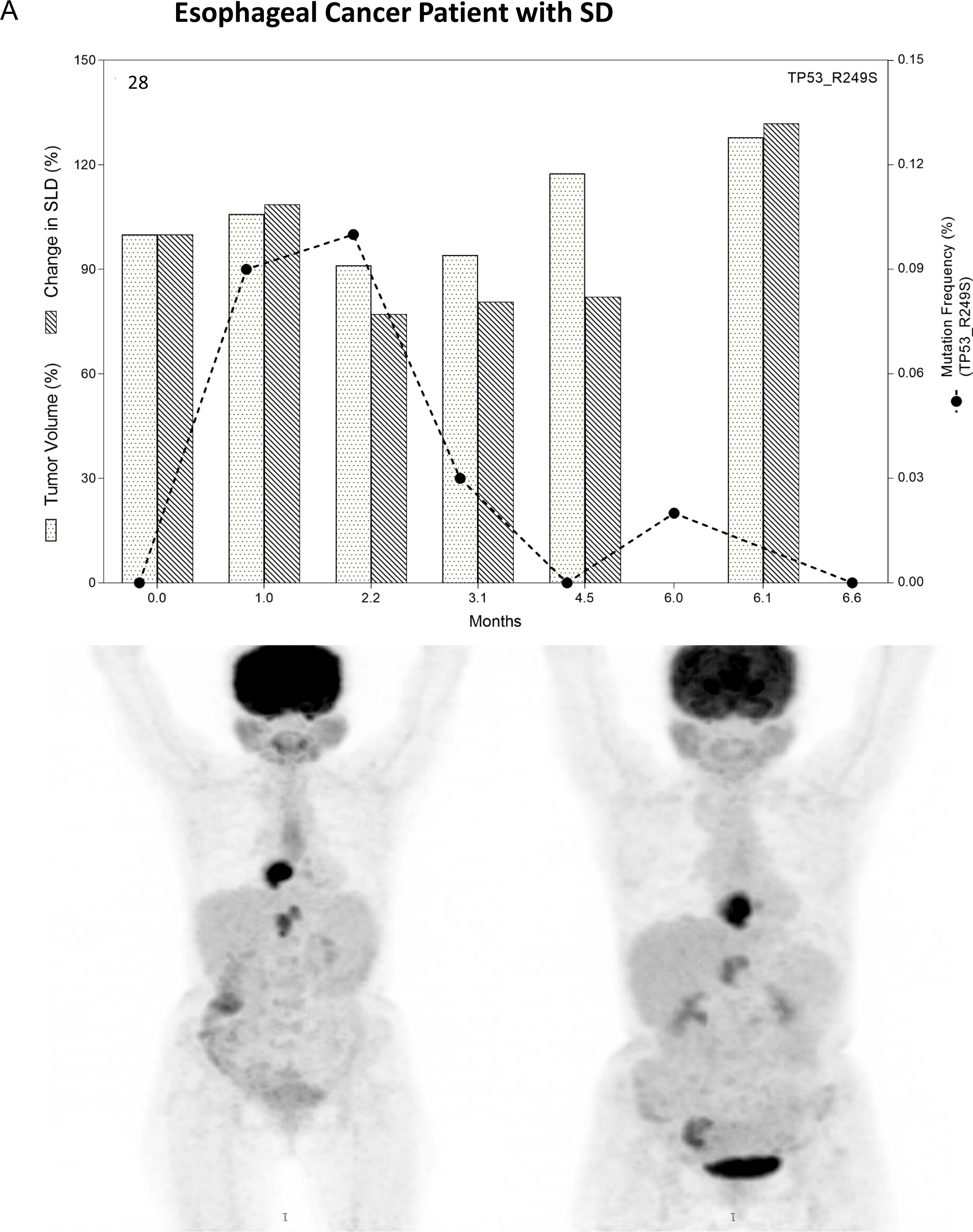
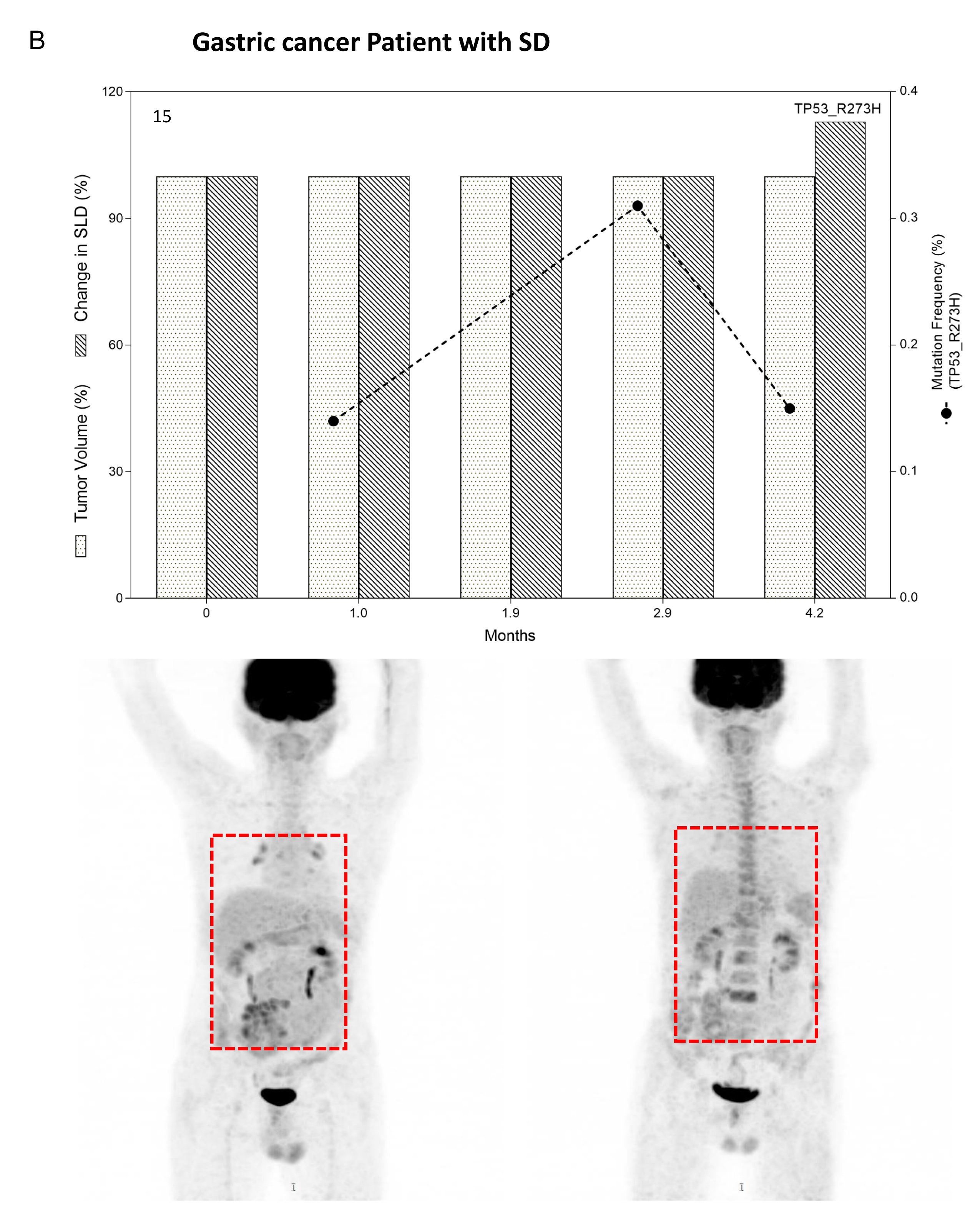
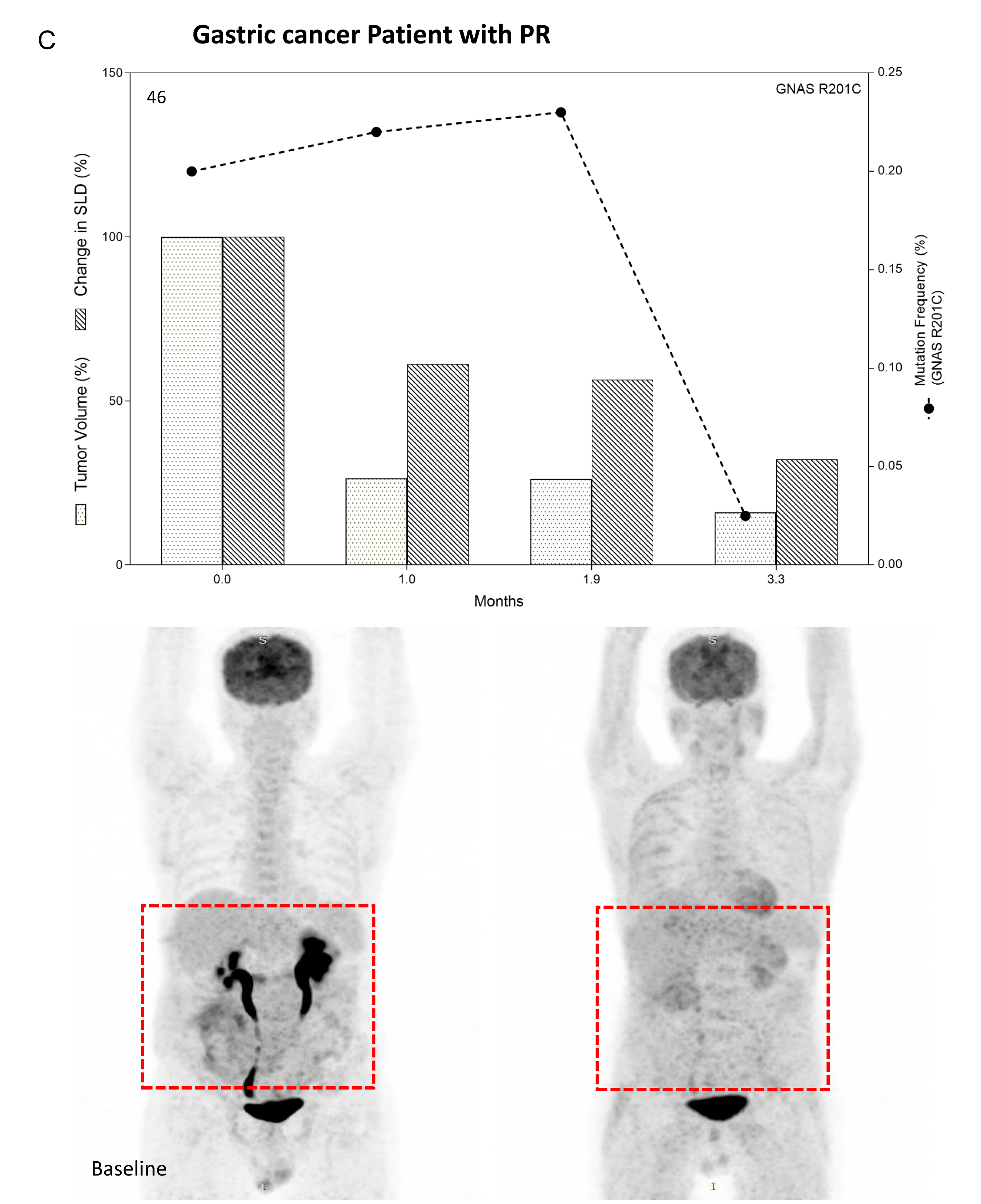
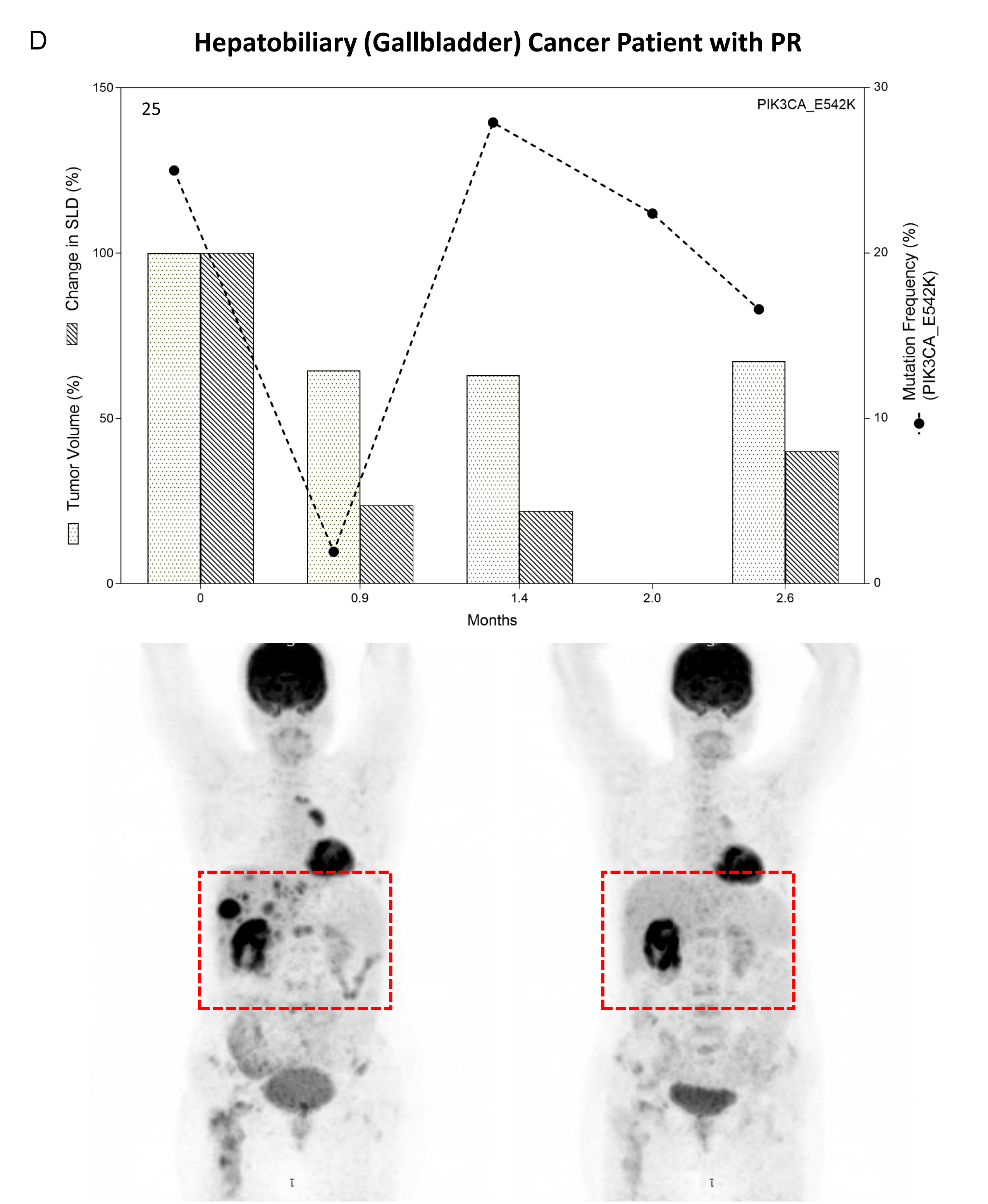
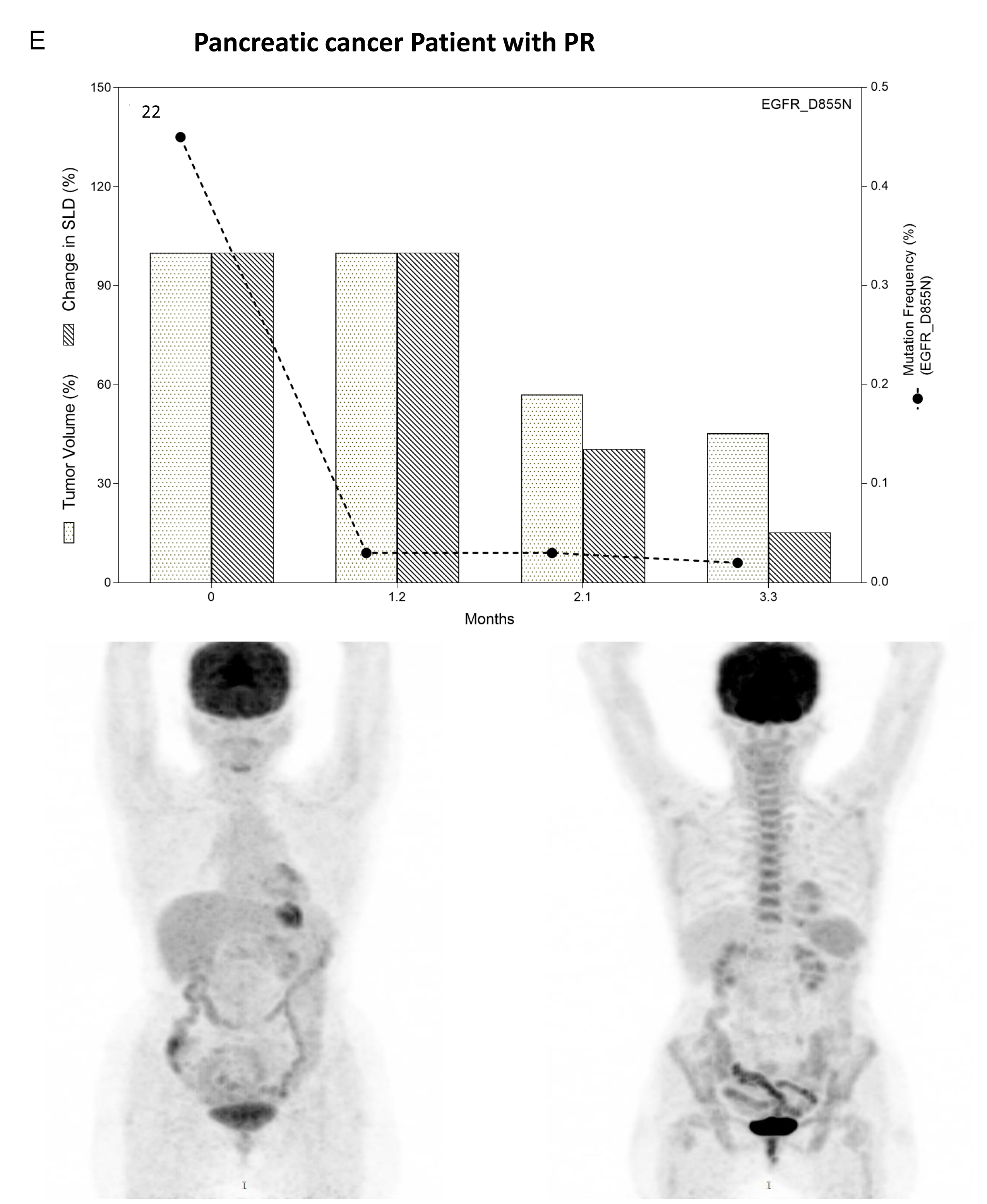
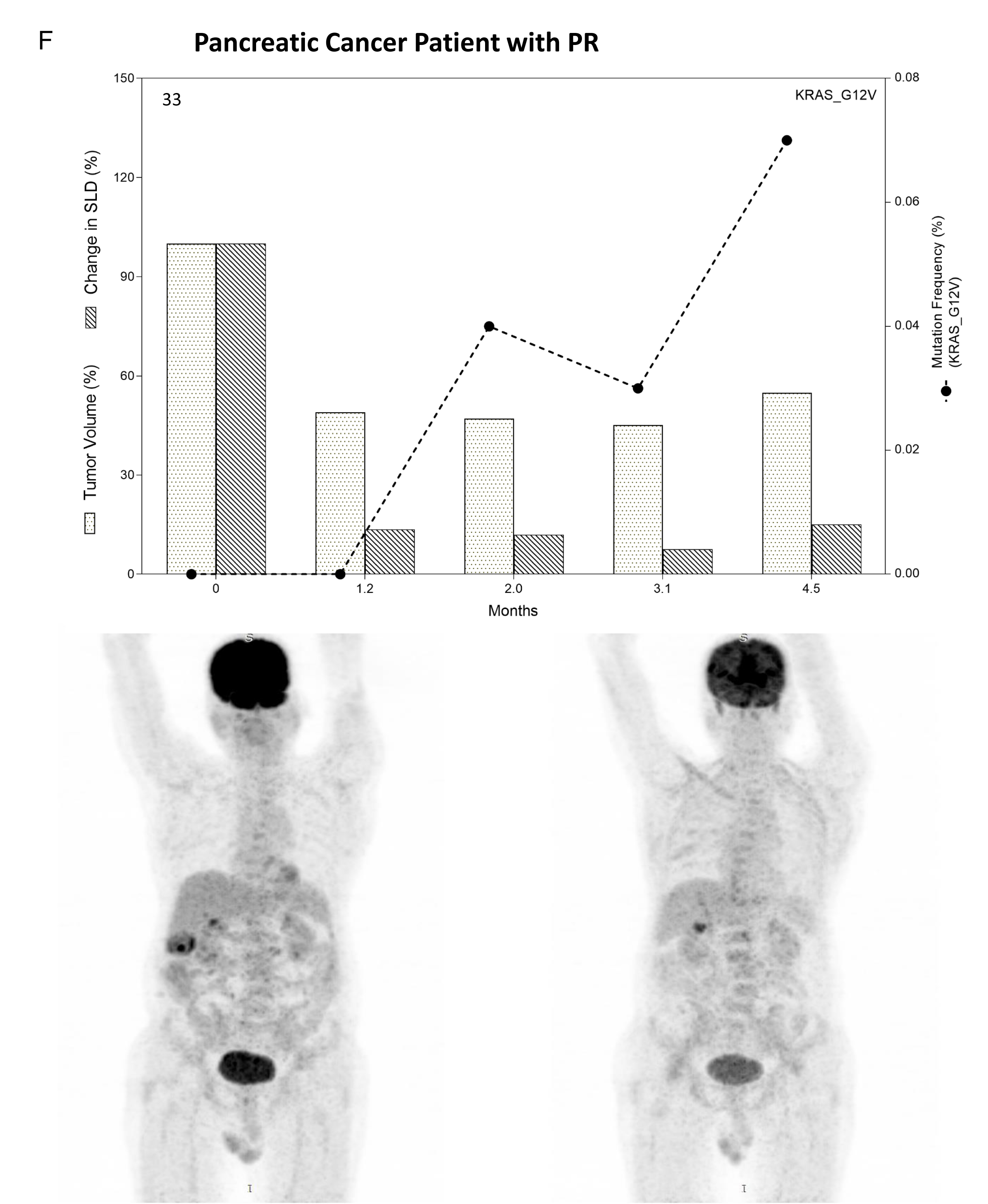
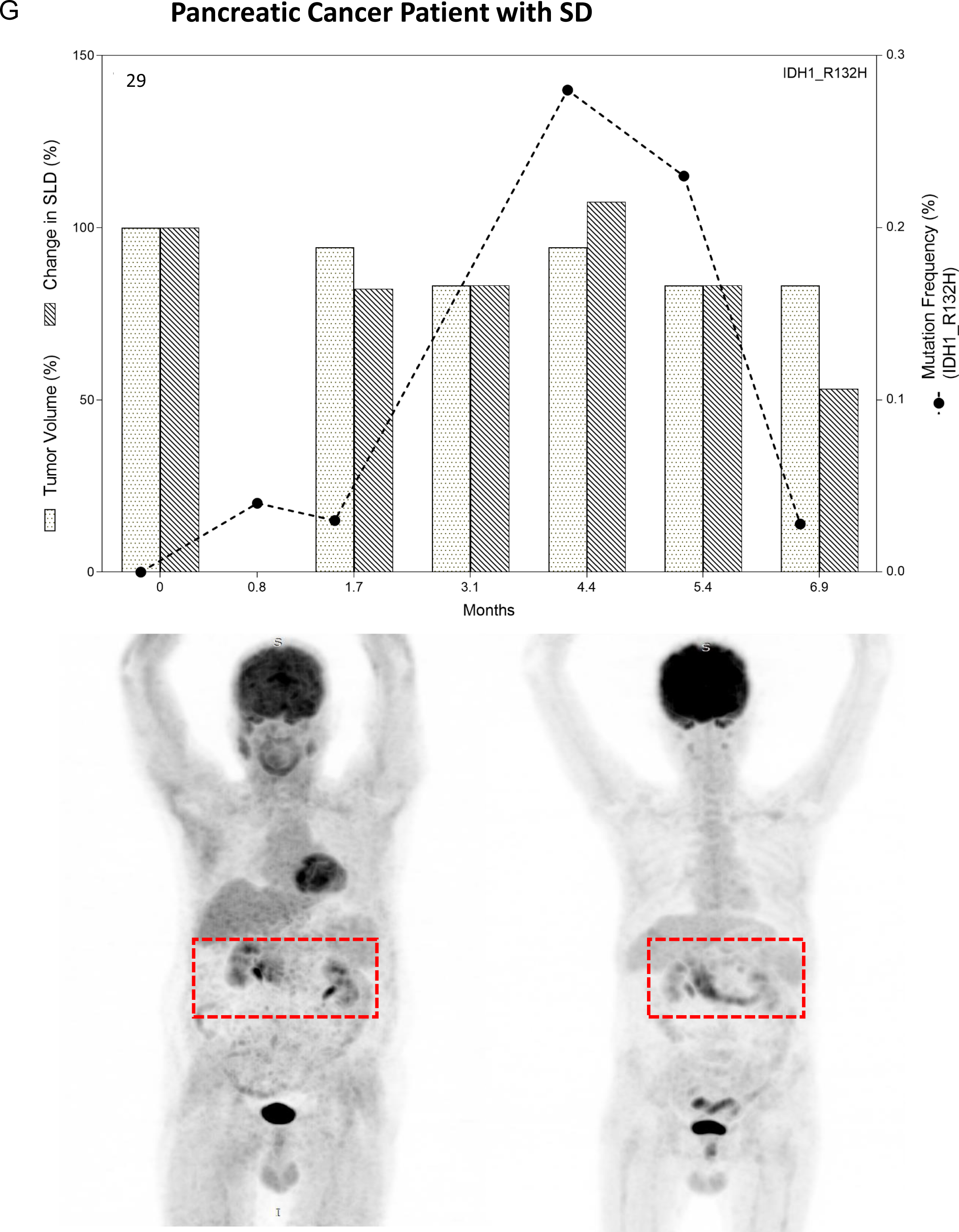
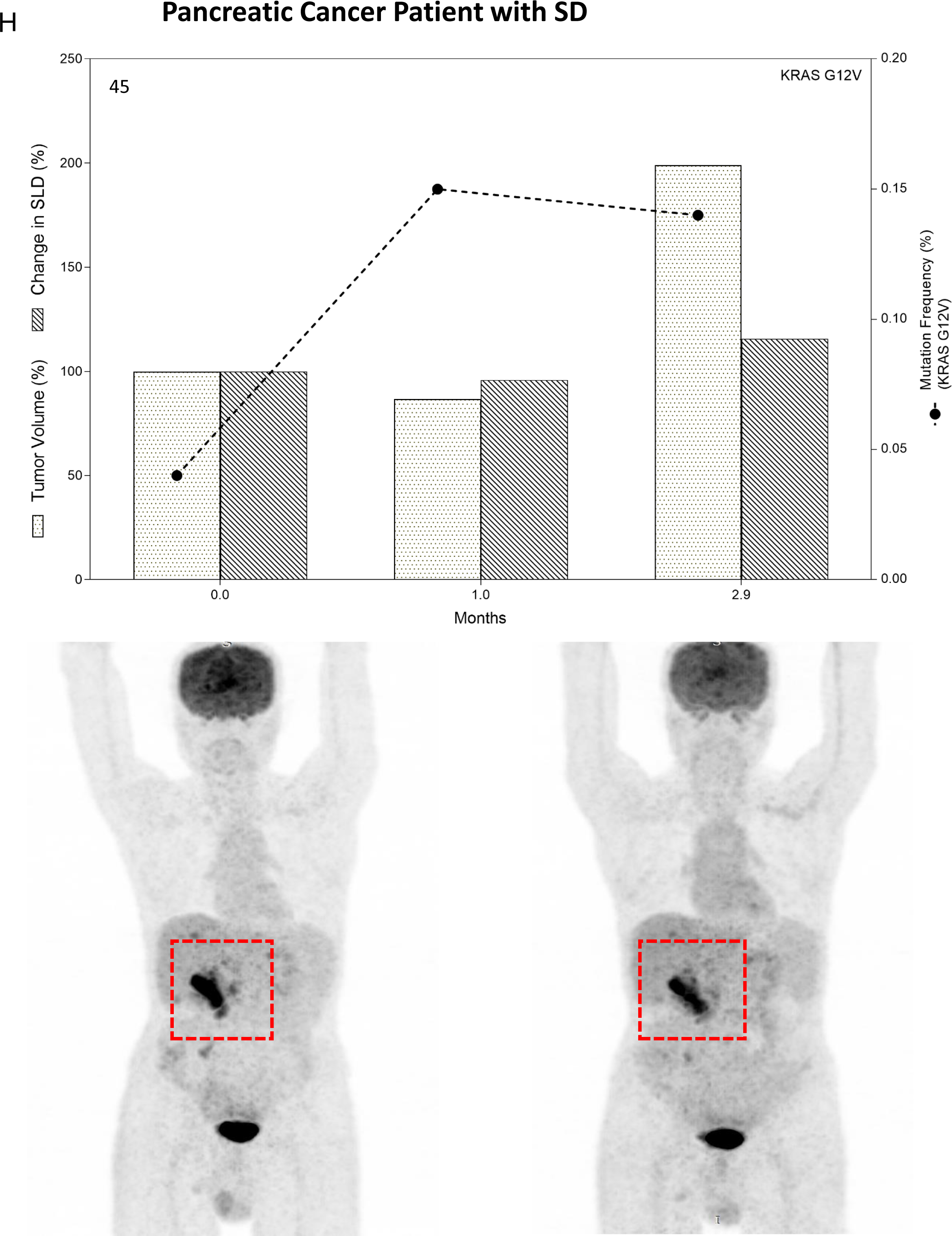
Monitoring Tumor Response by analyzing patient specific hot spot mutations by ddPCR in Esophageal (A), gastric (B-C), Hepatobiliary (D), and Pancreatic cancer patients (E-H), as mentioned in figure 3.

The role of ctDNA in monitoring treatment response was also evaluated in 1 hepatobiliary (gallbladder cancer) and 4 pancreatic cancer patients. In a case with hepatobiliary cancer (patient where we were able to evaluate tumor response just for 3 months after therapy, PIK3CA E542K MAF decreased along with tumor volume within 30 days of therapy, increased at 1.4 months follow-up and showed decreased trend thereafter (Figure 5D) with final outcome of PR.

Among 4 pancreatic cancer patients, 2 were with PR and 2 with SD. Patient 22 was monitored with EGFR D855N mutation in ctDNA which was consistently decreased according to tumor response to therapy (Figure 5E). Patient 33 was with PR till 5 months. In this case, KRAS G12V mutation was detected in tumor tissue at 24% MAF but initially undetected in ctDNA. About 2 - 5 months after therapy, MAF increased from 0.03% to 0.07% (Figure 5F). Additionally, two pancreatic cancer cases (patients 29 and 45) were with SD as determined by PET-CT and concurrent ctDNA based evaluation showed transient changes in the MAF (Figure 5G-H).

Therapy response was evaluated in four ovarian cancer patients. The patient with ovarian serous cystadenocarcinoma (patient 1) was monitored for 10 months after therapy administration and showed PR. Mutant allele TP53 R248W detection was concordant between tumor tissue and ctDNA and used to evaluate tumor burden which was consistently decreased in accordance with the decline in MAF (Figure 6A). Notably in this patient, ctDNA concentration, CA marker and MAF were positively correlated with tumor volume (r=0.7, p=0.1, r=0.84, p=0.0006 and r=0.96, p=0.3, respectively) (Table 2). Similarly, patient 16 was with PR till 5.7 months after receiving therapy and TP53 I254N MAF remained in the range of 1 to 1.7%. Notably, the MAF increased to 3.5% and continued to increase ∼8 and 11 months after therapy for which tumor size data was not available but change in MAF implicated PD (Figure 6B). Only 2 months follow-up after therapy for patient 2 indicated SD without notable change in MAF of TP53 K132E (Figure 6C). BRCA2 mutation (BRCA2 R2784W) positive ovarian cancer patient (patient 4) was exceptional where MAF was not in accordance with the disease progression (Supplementary Figure S2A). Single case of testicular cancer (patient 32) demonstrated remarkable tumor response with decreased tumor burden accompanied by decreased MAF of KRAS G12A (Figure 6D) which was positively correlated (r=0.96, p=0.6) (Table 2).

**Figure 6.**
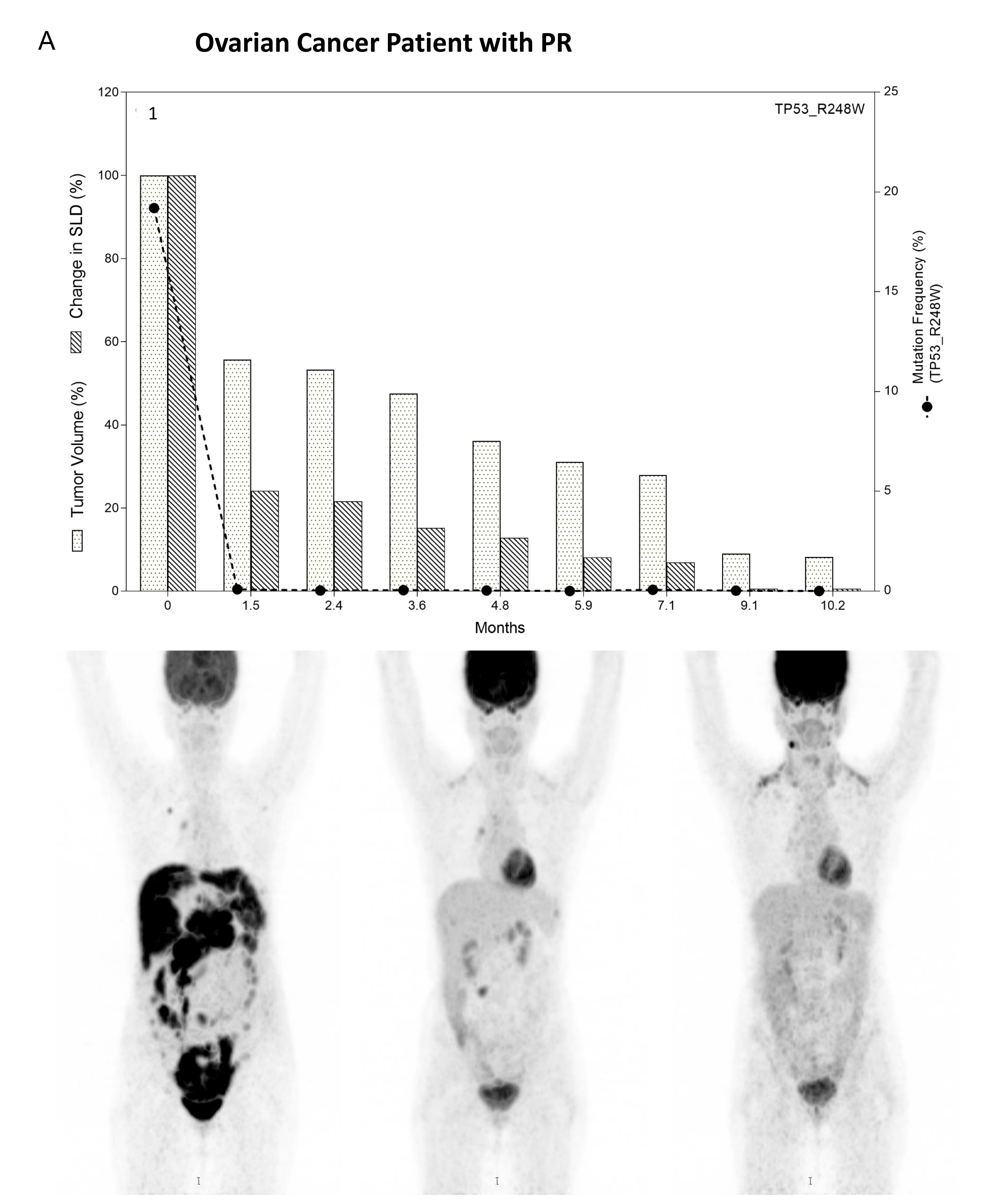
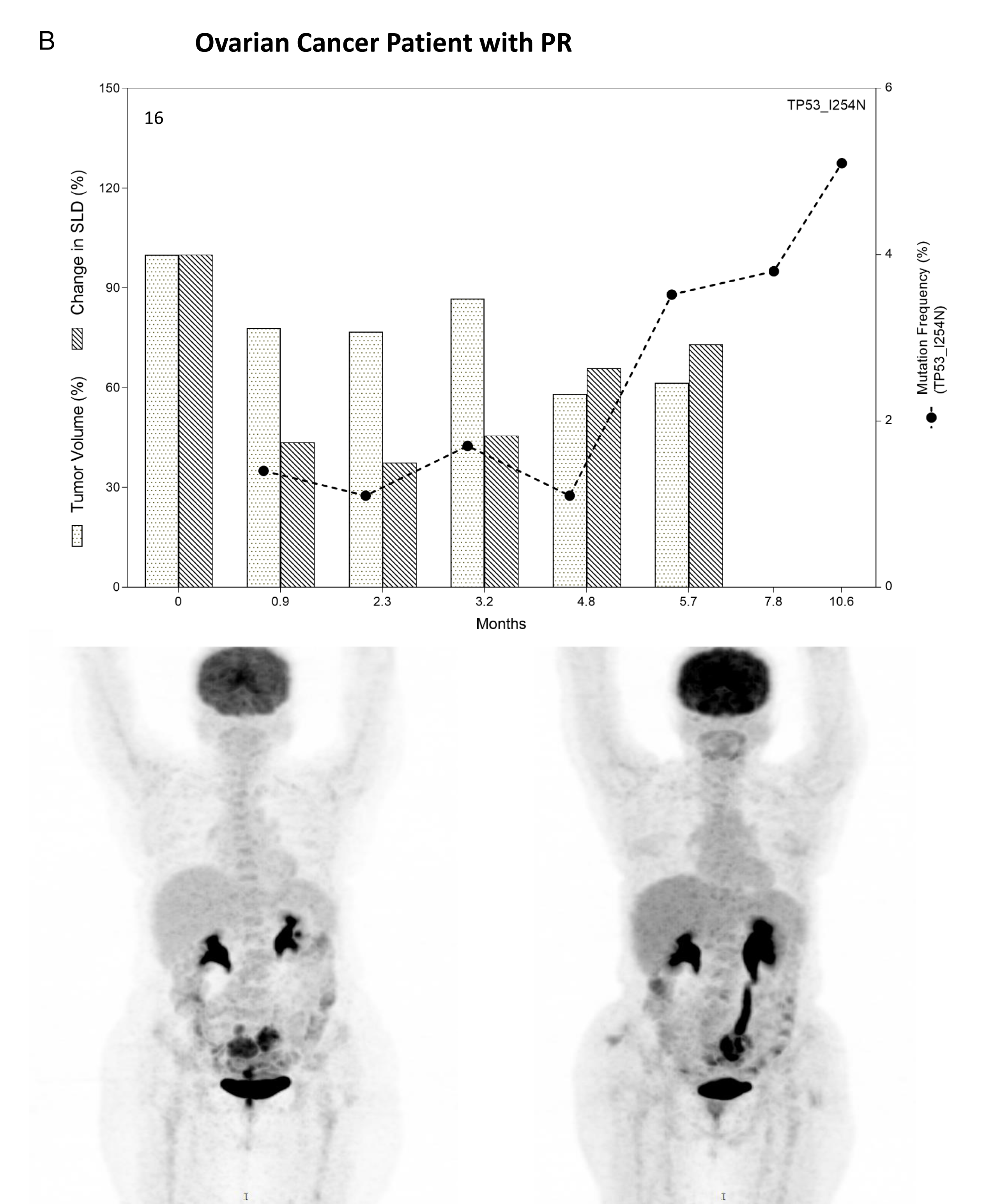
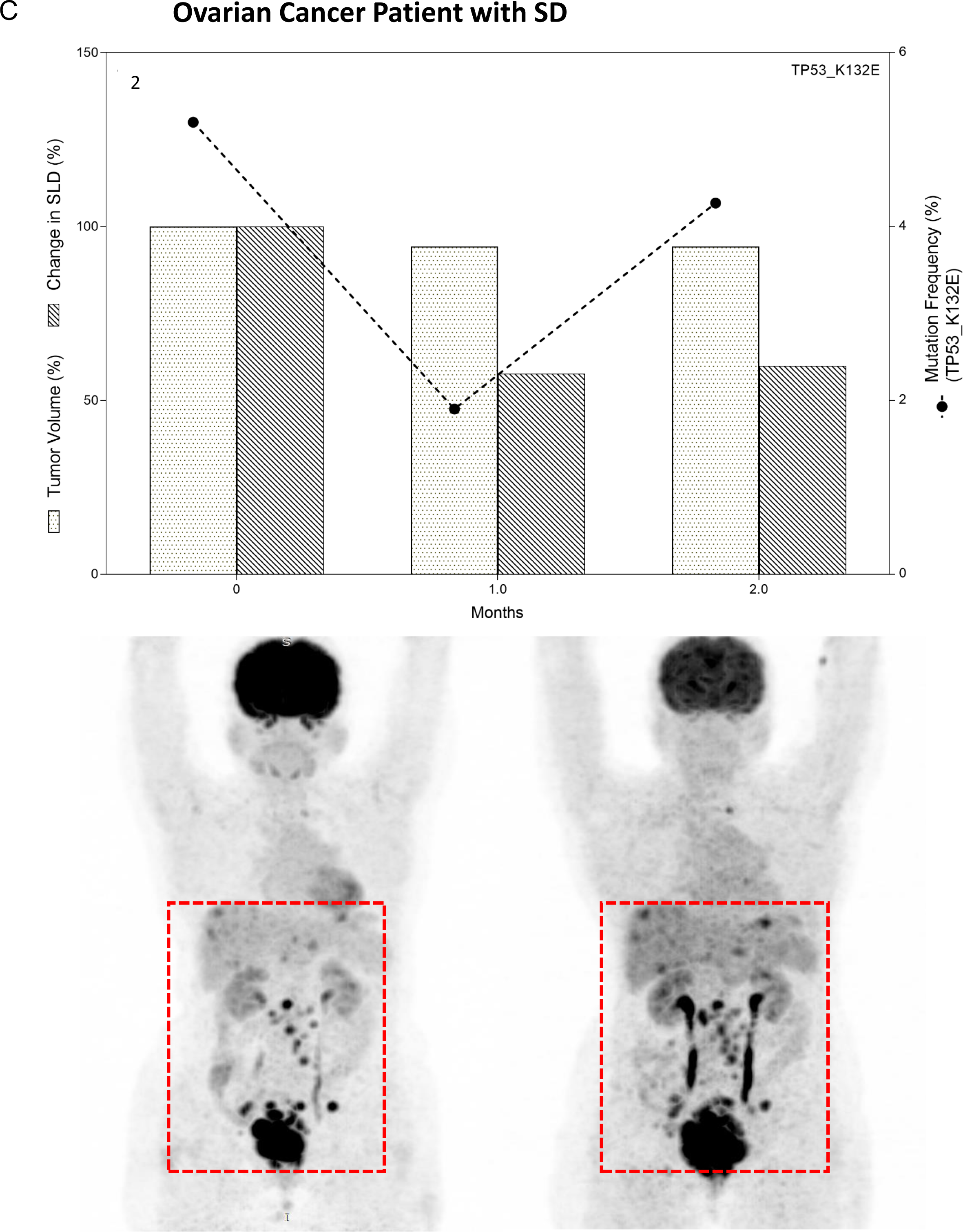
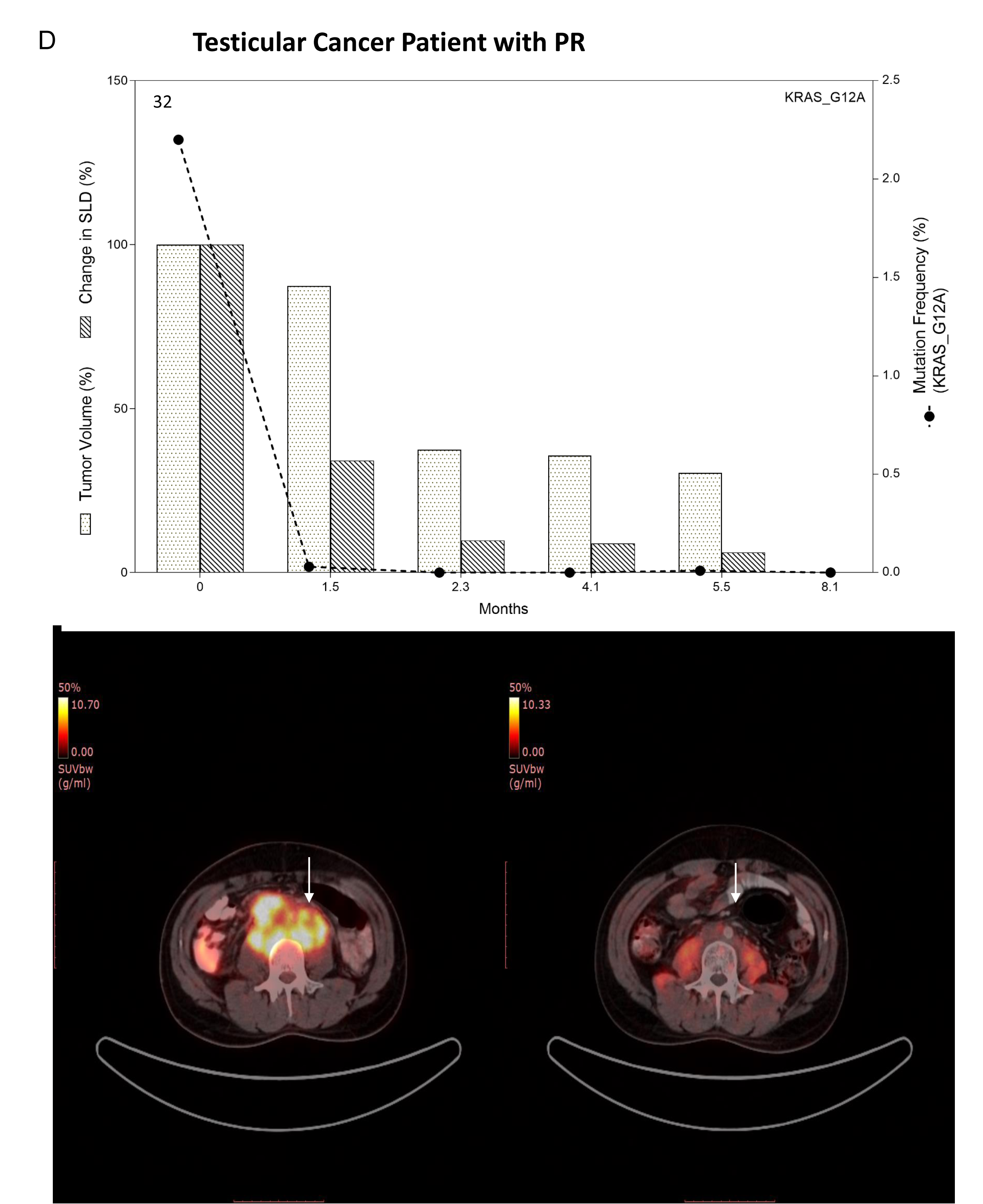
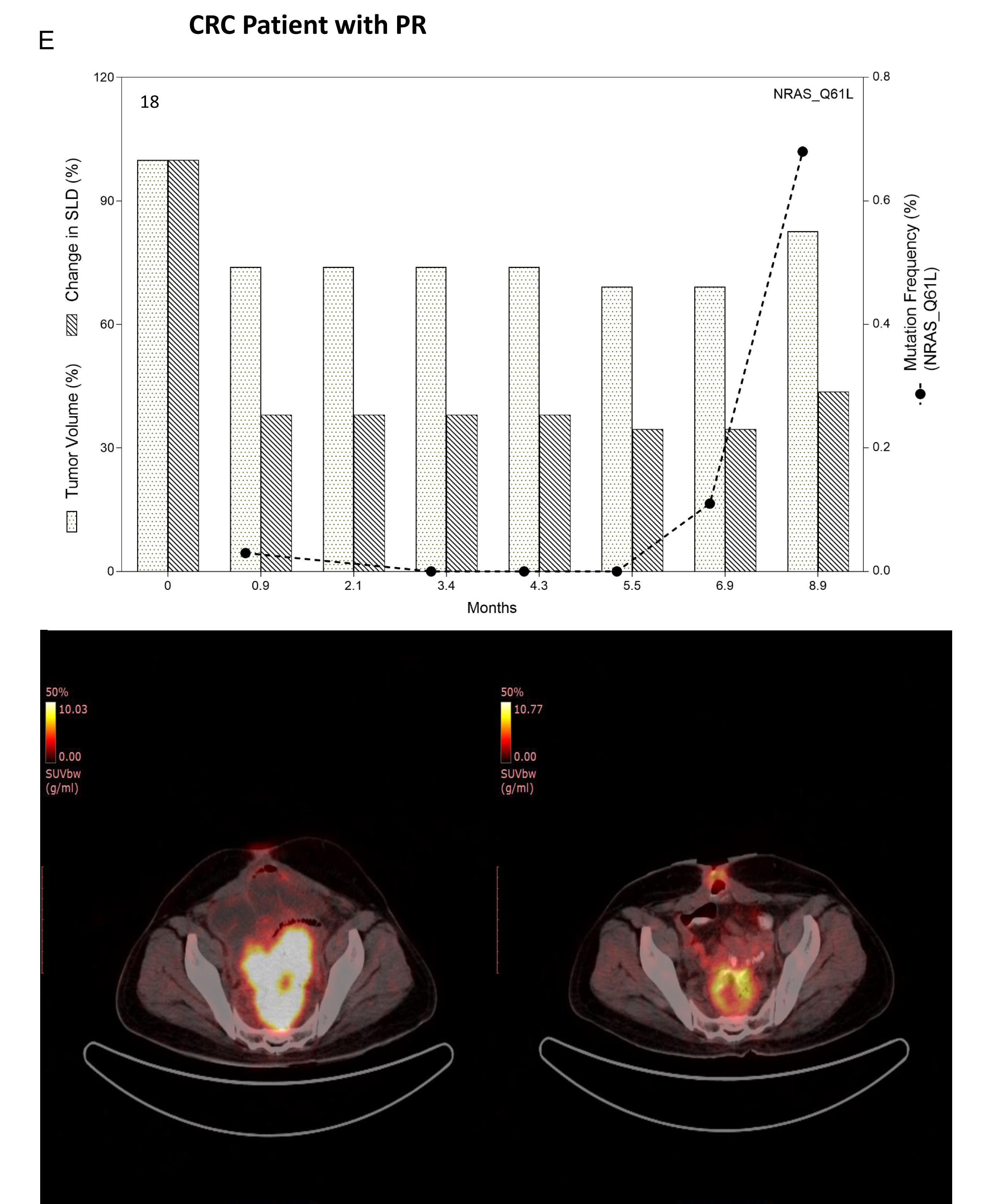
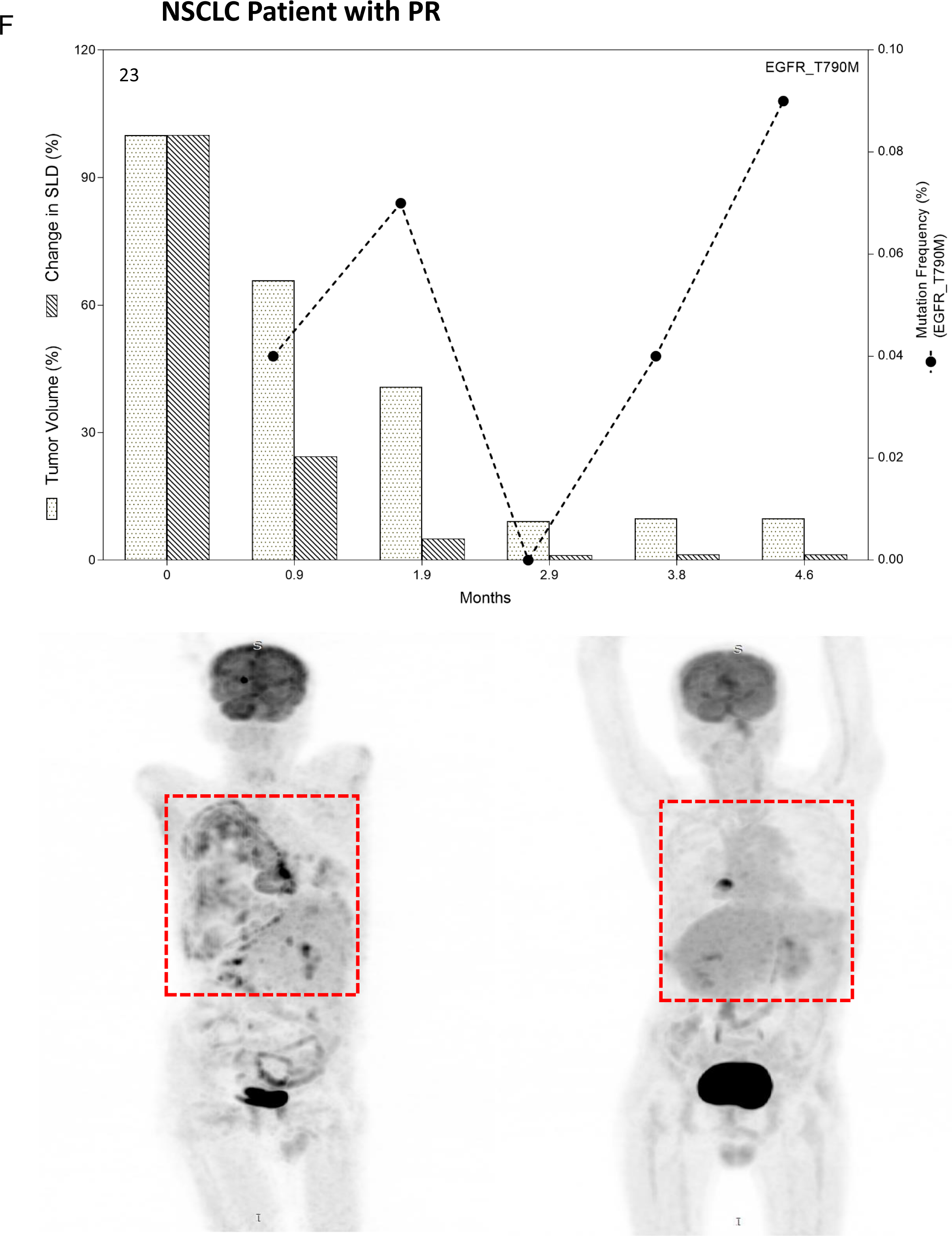
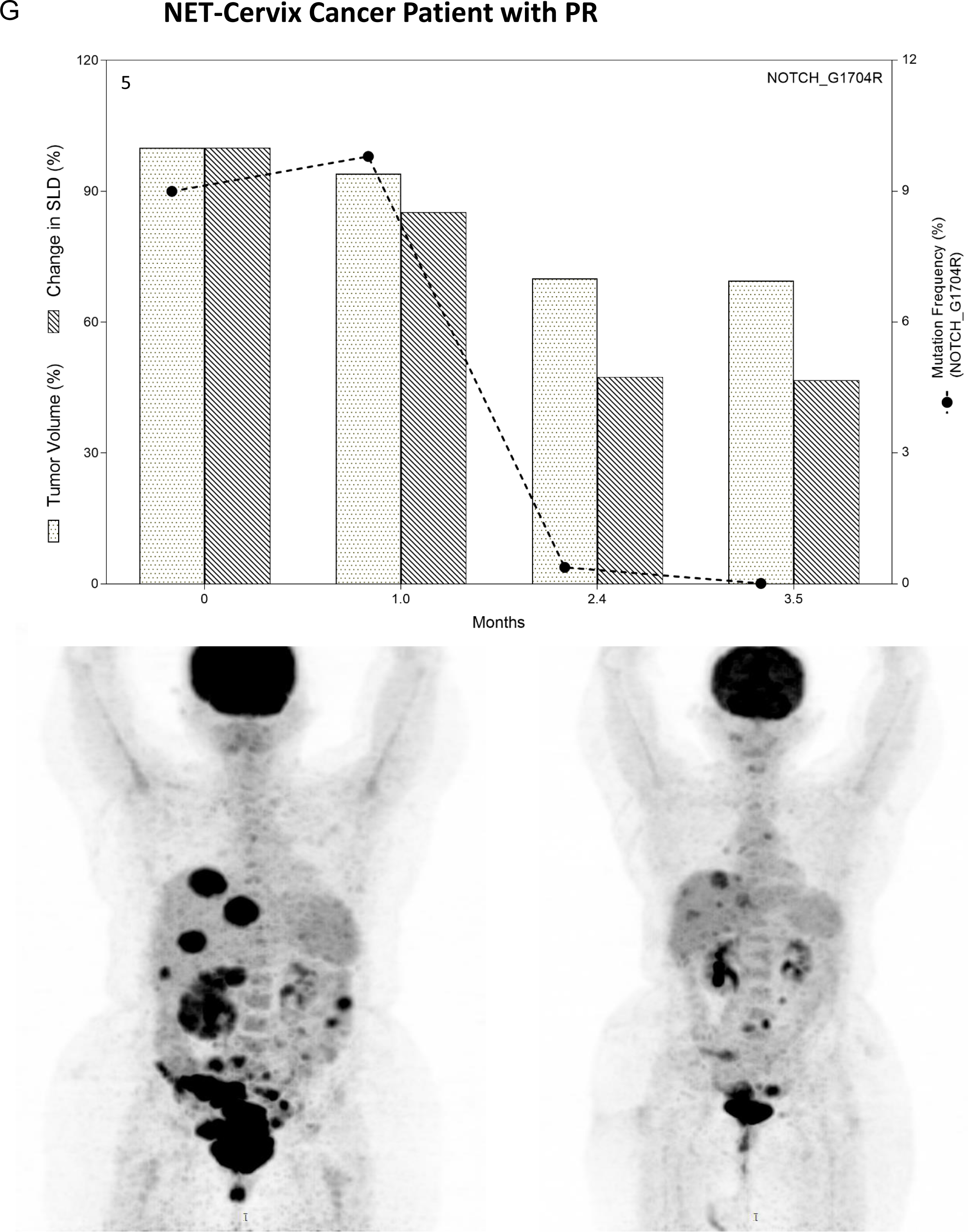
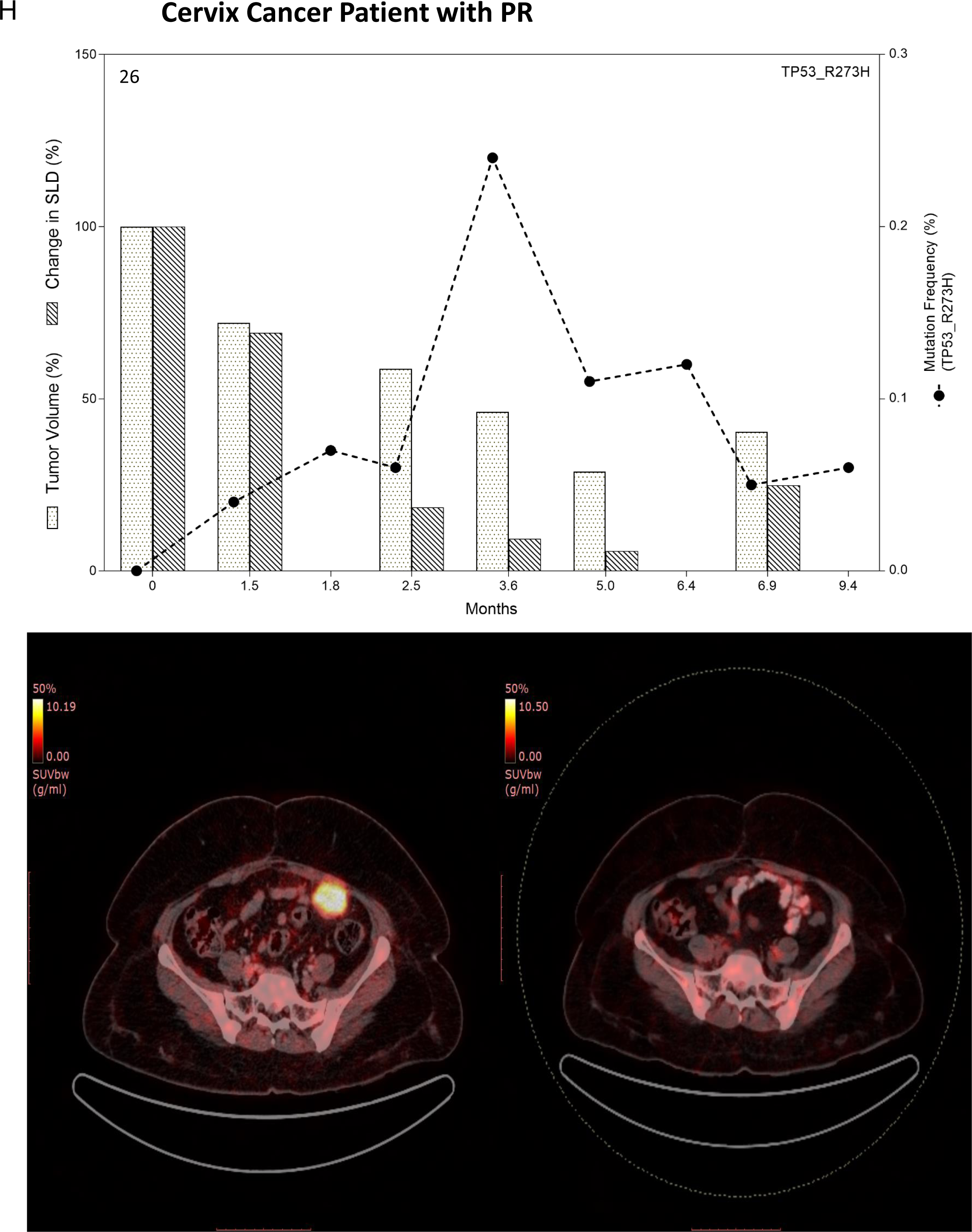
Monitoring Tumor Response by analyzing patient specific hot spot mutations by ddPCR in Ovarian (A-C), Testicular (D), Colorectal (E), NSCLC (F), and Cervical (G-H) cancer patients as mentioned in figure 3.

Among the six CRC patients monitored, patient 18 was evaluated with PR with concomitant decrease in NRAS Q61L MAF. However, increased trend of MAF in 7 and 9 months was in accordance with slight increase in tumor volume (Figure 6E). Four patients remained stable after therapy with subsequent changes in MAF of respective mutations (Supplementary Figure 2B-E). Interestingly, ctDNA of patient 10 harbored two driver mutations JAK2 V617F (1.6% MAF) and TP53 R175H (5.5% MAF) which were monitored during treatment response. Patient responded to therapy till 2.5 months and MAF of both mutant alleles remained quite stable. However, at 4 months post therapy, tumor burden started to increase with increased MAF of TP53 R175H (40.2%) in ctDNA while MAF of JAK2 V617F remained stable (0.9%) (Figure 6E). Remaining one patient (patient 6) showed disease progression and MAF of KRAS G12A remained unchanged 3.1-3.5% after 2 months of therapy (Supplementary Figure S2F).

Two lung cancer cases were with SD and 2 with PR. Lung adenocarcinoma patient 23 who was previously treated with Afatinib and Erlotinib before enrolment in the study was positive for both EGFR T790M and exon 19 (L747-P753delinsS) deletion mutations. He received Osimertinib along with combination of other systemic agents which resulted in PR with transient increase in MAF of EGFR T790M from 0.04 to 0.09% by ddPCR (Figure 6F). NSCLC patient 7 showed transitions of SD and PD with similar trend in MAF during 11 months follow-up (Supplementary Figure S2G). Similarly, the third lung adenocarcinoma patient (patient 40) receiving Afatinib (EGFR L858R mutation) and Everolimus (PTEN mutation) along with Vinorelbine was positive for resistance mutation T790M in ctDNA. This lead to replacement of Afatinib with Osimertinib. This further continued SD in patient till last follow-up (Supplementary Figure S2H).

Monitoring tumor response in three cervical cancer patients revealed two patients (patient 5 and 26) with PR (Figure 6G-H) and 1 with SD (patient 13) as determined by limited follow-up of tumor volume and respective MAF in ctDNA (Supplementary Figure S2I).

### Correlation between the level of mutated ctDNA, CA markers, ctDNA concentration and tumor burden

ctDNA quantification, determination of MAF by ddPCR, and CA markers was successful in 40 patients with >3 longitudinal time points. We evaluated the correlation of all these liquid biopsy analytes with tumor burden during response assessment of each patient. Interestingly, MAF was found to be mostly correlated with tumor burden in 16/19 (85%) patients with PR and overall in 27/40 (68%) patients. Among these, there was significant correlation in 13 (48%) patients with P-value <0.05 (Table 2) while in 14 patients (52%) there was positive correlation between MAF and tumor burden with P-value ranged from 0.06 to 0.6 (Table 2). This was followed by positive correlation of CA markers (n=19 patients, 47.5%), ctDNA concentration (per ml of plasma) (n=18 patients, 45%) with tumor response (Table 2).

### Association of ctDNA concentration with tumor metastasis and Clinical response

We compared the baseline ctDNA concentrations from patients (n=60) with different cancer types included in this cohort with ctDNA from healthy individuals (n=20) (1 patient specimen each of testicular, prostate, medulloblastoma and yolk sac tumor were excluded from this analysis due to unavailability of multiple patients of respective cancer types). ctDNA yield varied among different tumor types but was significantly higher compared to ctDNA yield from healthy subjects (Figure 7A). Increase in ctDNA from 4 ovarian cancer patients was not significant probably due to limited sample size. Next, we compared the ctDNA concentration (ng/mL of plasma) between patients with single metastatic site (n=25) and patients with 2-5 metastatic sites (n=31). Baseline ctDNA concentration was considered for this analysis. Median ctDNA concentration in patients with single metastatic site was 15.6 ng/mL (range 2 to 48 ng/mL) and in patients with 2 -5 metastatic sites was 24.6 ng/mL (range 11 to 184 ng/mL). Notably, ctDNA concentration was higher in patients with 2-5 metastatic sites (*P=0.04*) compared to single metastatic site (Figure 7B).

**Figure 7.**
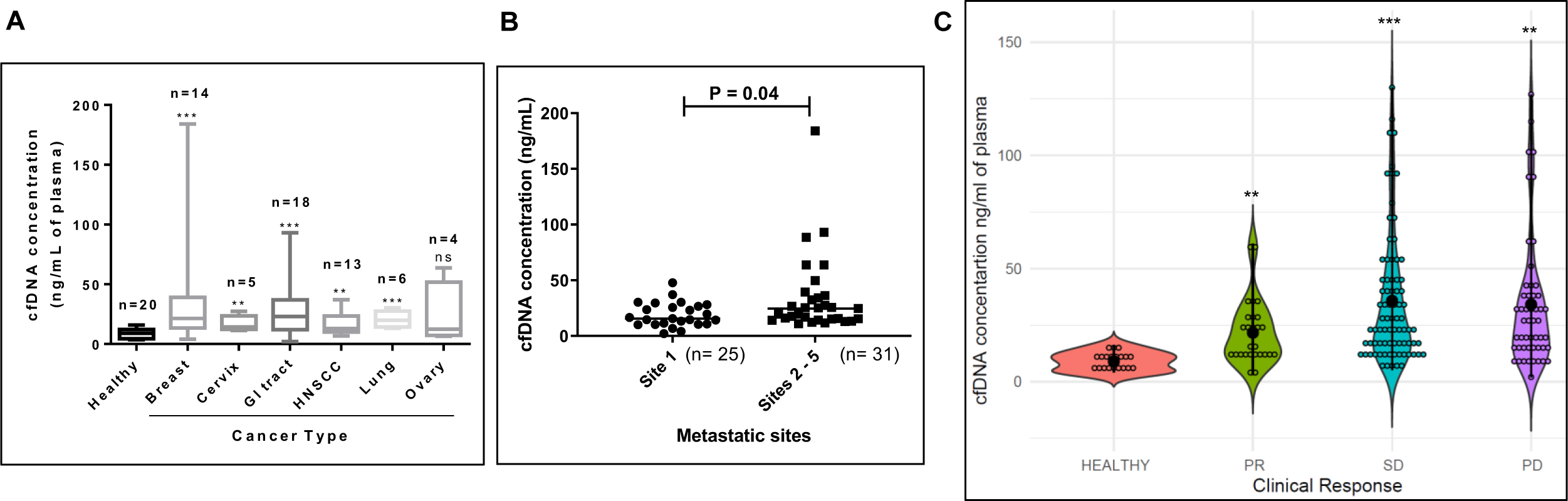
Association of circulating free DNA (ctDNA) concentrations (ng/mL of plasma) with tumor metastasis and clinical response. (A) Baseline ctDNA yield in different malignancies compared with ctDNA from healthy subjects. (B) Compared the ctDNA concentrations between patients with single metastatic site (n=25) and 2 -5 metastatic sites (n =31). (C) Violin plot of ctDNA concentrations from healthy individuals (n=20), and different clinical response groups; PR (n=27 measurements), SD (n=80 measurements) and PD (n=51 measurements). Black dots represented the group mean in each category (** P < 0.01 and *** P < 0.001).

Furthermore, we assessed the ctDNA levels among healthy individuals (n=20) and different clinical response groups; PR (n=27 measurements), SD (n=80 measurements) and PD (n=51 measurements). Median ctDNA levels were significantly higher in patients with PR (16 ng/mL, range 3- 61), SD (25 ng/mL, range 5 -130) and PD (26 ng/mL, range 2 -127) than those in healthy counterparts (9 ng/mL, range 4-16) (Figure 7C). Among the clinical response groups, ctDNA levels from patients with SD and PD were significantly higher (*P = 0.01* and *P = 0.04*, respectively) compared to PR group (Figure 7C).

## Discussion

Here, we report the retrospective analysis of cancer patients from RESILIENT trial ^15^ across 11 different cancer types to reveal the potential of blood-based biomarkers (ctDNA, MAF and CA markers) in assessing the tumor response following RECIST criteria to the therapy guided by ETA analysis. A total of 48 patients were evaluated longitudinally for this analysis (Figure 1, Table 1).

In this study, clinical utility of cell free tumor DNA was evaluated by patient-specific ctMRD assays longitudinally and correlated with tumor volume (cm^3^) as well as RECIST criteria guided response assessment. Among 40 patients monitored, 19 patients (47.5%) showed PR, 18 patients (45%) showed SD and 3 (7.5%) showed PD (Table 2). Longitudinal analysis showed that ctDNA dynamics in allele frequency was consistent with therapy response in all 40 patients evaluated. MAF correlated with tumor volume in 27/40 (68%) patients. Maximum correlation of MAF with tumor response was found for 16/19 patients with PR (84%) (Table 2). In comparison, as the standard clinical biomarkers, cancer antigens correlated with tumor burden in only 19/40 (47.5%) cases highlighting their limited utility for tumor monitoring in the study population. Therefore, ctDNA as a biomarker was found to be superior to conventional serum biomarker in tracking therapy response similar to other studies that also found superiority of ctDNA over standard biomarkers ^18, 19^. Recently, we utilized ctDNA based liquid biopsy approach to serially evaluate mutations in ctDNA to monitor disease status, ascertain treatment response, identify emergent drug resistance and detect recurrence at sub-radiological levels in a case of metastatic non-small cell lung cancer (NSCLC), with prior history of treated colorectal cancer as well as in periampullary carcinoma ^20, 21^.

The importance of appropriate selection of driver mutant allele to monitor tumor response was seen in colorectal cancer patient 10. This patient’s ctDNA harbored two driver mutations JAK2 V617F (1.6% MAF) and TP53 R175H (5.5% MAF) which were monitored during treatment response. Patient responded to therapy till 2.5 months and MAF of both mutant alleles remained quite stable. However, at 4 months post therapy, tumor burden started to increase with increased MAF of TP53 R175H (40.2%) in ctDNA while MAF of JAK2 V617F remained stable (0.9%) (Supplementary Figure S2E). This also implicated presence of JAK2 V617H mutation in CRC patient despite the findings from Spanish study wherein JAK2 V617F mutation was not evident in 75 CRC patients ^22^. The importance of JAK2 V617F mutation in CRC development thus can be explored further. Similarly, monitoring tumor response with the help of specific EGFR alterations allowed us to monitor treatment responses in lung cancer patients. For example, the lung adenocarcinoma patient (patient 40) receiving Afatinib (EGFR L858R mutation) and Everolimus (PTEN mutation) along with Vinorelbine initially responded to therapy until the appearance of TKI resistance mutation T790M based on which Afatinib was replaced by Osimertinib (Supplementary Figure S2H). Additionally, the discovery of rare mutations were useful in monitoring, e.g., IDH1 R132H mutation in pancreatic adenocarcinoma patient (patient 29); in the sole case reported previously wherein the patient progressed after just 1 cycle of therapy ^23^. Similarly, pancreatic cancer patient 22 was positive for EGFR D855N, a point mutation in Exon 21; while the clinical significance was unknown, computational modeling predicted it as TKI resistant mutation ^24^. Future studies are required to better understand the role of IDH1 and EGFR mutations in PDA.

The total quantity of cell free DNA was previously reported to show correlation with tumor volume ^19, 25^. In this study we show that quantitative cfDNA correlated with tumor volume in only 44% cases (Table 2). Since the majority of cfDNA originates from non-malignant cells ^26^, and tumor cells contribute to a minor proportion of this cfDNA, total cfDNA measurements are neither indicative of tumor burden nor treatment response. Notably, higher plasma cfDNA was observed in cancer patients compared to healthy individuals (Figure 7A) which was significantly associated with number of metastatic sites in the cancer individuals compared to single metastatic site implicating the ability of cfDNA to predict possibility of having multiple metastasis of the tumor ^27^. The patient cohort in our study did not have any patients with localized (non-metastatic) cancer to compare cfDNA concentration with. Total plasma cfDNA levels were higher in the cancer cohort (than healthy individuals) irrespective of clinical response groups (PR, SD and PD) (Figure 7C). Median cfDNA levels were higher in patients with SD and PD compared with PR which is in agreement with the previous reports where a decline in post-treatment plasma DNA levels has been observed in responders ^28, 29^. These findings suggest that monitoring the levels of plasma cfDNA during the course of cytotoxic / targeted therapies may help in predicting response to therapy.

Monitoring of disease response carries enormous importance to check the progress on the disease course during the active treatment phase as well as post-treatment surveillance period. This is currently achieved by monitoring symptoms, radiological imaging and measurements of serum tumor markers. Risk of radiation exposure and inconvenience limits the frequency with which radiological scans can be performed. Therefore, radiological assessments are usually not conducted before completion of therapy unless patient experiences clinical symptoms of disease progression. Analysis of major driver mutations using ctMRD based liquid biopsy analysis could be performed to explore precise individualized treatments. With the advancement of ctDNA technology and sensitive detection platforms such as ddPCR, this non-invasive non-radiological method, which supports longitudinal evaluations has the potential to guide therapeutic decisions including course corrections based on real time monitoring of disease status / treatment responses. We believe that screening for driver mutations via tumor and plasma multigene sequencing can guide selection of patient-specific ctMRD assays to monitor disease status and treatment response in a personalized manner with high sensitivity and specificity with patient specific baselines.

## Funding

No external funding was received for the present study.

## Data Availability

The minimal data set underlying the results described in the manuscript are within the manuscript and its Supporting Information file.

## Acknowledgements

The authors express gratitude towards the patients as well as families of patients who consented to participate in the RESILIENT study; the staff at HCG-MCC for patient care and management.

## Author Contributions

Conceptualization, Dadasaheb Akolkar, Darshana Patil, Sachin Apurva, Navin Shrivastava and Rajan Datar; Data curation, Sneha Puranik, Swapnil Puranik, Sachin Apurva, Harshal Bodke, Jinumary John and Ajay Srinivasan; Formal analysis, Dadasaheb Akolkar, Darshana Patil, Sneha Puranik, Swapnil Puranik, Pratiksha Nandre, Karishma Sabale, Sachin Apurva, Harshal Bodke, Jinumary John and Ajay Srinivasan; Funding acquisition, Rajan Datar; Investigation, Dadasaheb Akolkar, Sneha Puranik, Swapnil Puranik, Pratiksha Nandre, Karishma Sabale, Navin Shrivastava and Ajay Srinivasan; Methodology, Dadasaheb Akolkar, Sneha Puranik, Swapnil Puranik, Navin Shrivastava and Ajay Srinivasan; Project administration, Dadasaheb Akolkar, Darshana Patil, Sachin Apurva, Navin Shrivastava, Vineet Datta, Stefan Schuster and Rajan Datar; Resources, Dadasaheb Akolkar, Darshana Patil, Navin Shrivastava, Vineet Datta, Stefan Schuster and Rajan Datar; Software, Sachin Apurva, Harshal Bodke and Ajay Srinivasan; Supervision, Dadasaheb Akolkar, Darshana Patil, Sneha Puranik, Swapnil Puranik, Sachin Apurva, Navin Shrivastava and Rajan Datar; Validation, Dadasaheb Akolkar, Sneha Puranik, Swapnil Puranik, Pratiksha Nandre, Karishma Sabale, Sachin Apurva, Harshal Bodke and Navin Shrivastava; Visualization, Dadasaheb Akolkar, Darshana Patil, Sachin Apurva, Harshal Bodke, Navin Shrivastava, Vineet Datta, Ajay Srinivasan and Rajan Datar; Writing – original draft, Dadasaheb Akolkar, Jinumary John and Ajay Srinivasan; Writing – review & editing, Dadasaheb Akolkar, Darshana Patil, Sneha Puranik, Swapnil Puranik, Pratiksha Nandre, Karishma Sabale, Sachin Apurva, Harshal Bodke, Navin Shrivastava, Vineet Datta, Jinumary John, Ajay Srinivasan and Rajan Datar.

## Ethics approval and informed consent

All biological samples were obtained from patients in the RESILIENT trial (CTRI/2018/02/011808). The study was approved by the Ethics Committees of both the participating institutions - Datar Cancer Genetics and HCG Manavata Cancer Centre. All patients provided written informed consent. The study was performed in accordance with the Declaration of Helsinki.

## Conflict of Interest

All authors are employees of Datar Cancer Genetics, India which offers commercial services in the area of onco-diagnosis and therapy management.

## Supplementary figures and tables

Supplementary Figure S1. Annealing temperature optimization for representative custom made TaqMan assays (JAK2 V617F) (A) and IDH R132H (B). Red circled droplets shows false positive mutants detected in the wild-type allele specific assay and well highlighted with red rectangle shows optimum annealing temperature for the assay. (C) Limit of detection was performed for IDH1 R132H by spiking mutant only and wildtype DNA to obtain frequency of 0.5% (well D05), 0.05% (well C05), 0.01% (well B05), 0.005% (well E05) and well F05 contained only WT control and A05 contained NTC (Non-template control).

Supplementary Figure S2. Monitoring Tumor Response by analyzing patient specific hot spot mutations by ddPCR in representative disease progressed patients: Ovarian (A), Colorectal (B-F), NSCLC (G-H) and cervical cancer patients (I) as mentioned in figure 3. Representative coronal views from PET-CT Images of patients at Baseline and Follow-Up scan are shown in lower panel of respective graph. Tumor measurements were by RECIST 1.1 criteria (Single lesion dimension (SLD %) and ellipsoid tumor volume (cm^3^).

Supplementary Table S1 List of individual patient wise Taqman assays (ctMRD) used in ddPCR (Values in bold fonts represent concordant mutations in tumor tissue and plasma).

Supplementary Table S2 Optimization of ddPCR assays for annealing temperature along with suitable positive controls.

Supplementary Table S3. CA marker (cancer antigen) determination of clinically evaluable patients (n=48).

Supplementary Table S4. List of 452 genes analyzed by Next generation Sequencing (NGS).

Supplementary Table S5. List of 52 genes analyzed by Next generation Sequencing (NGS) using circulating tumor DNA (ctDNA)

## Notes

### Clinical Trial

CTRI/2018/02/011808

### Author Declarations

The study was approved by the Ethics Committees of both the participating institutions - Datar Cancer Genetics and HCG Manavata Cancer Centre. All patients provided written informed consent. The study was performed in accordance with the Declaration of Helsinki.

