## Supplementary figures and tables for "Evaluation of patient-specific cell free DNA assays for monitoring of minimal residual disease in solid tumors": Supplementary figures.pptx

### Slide 1
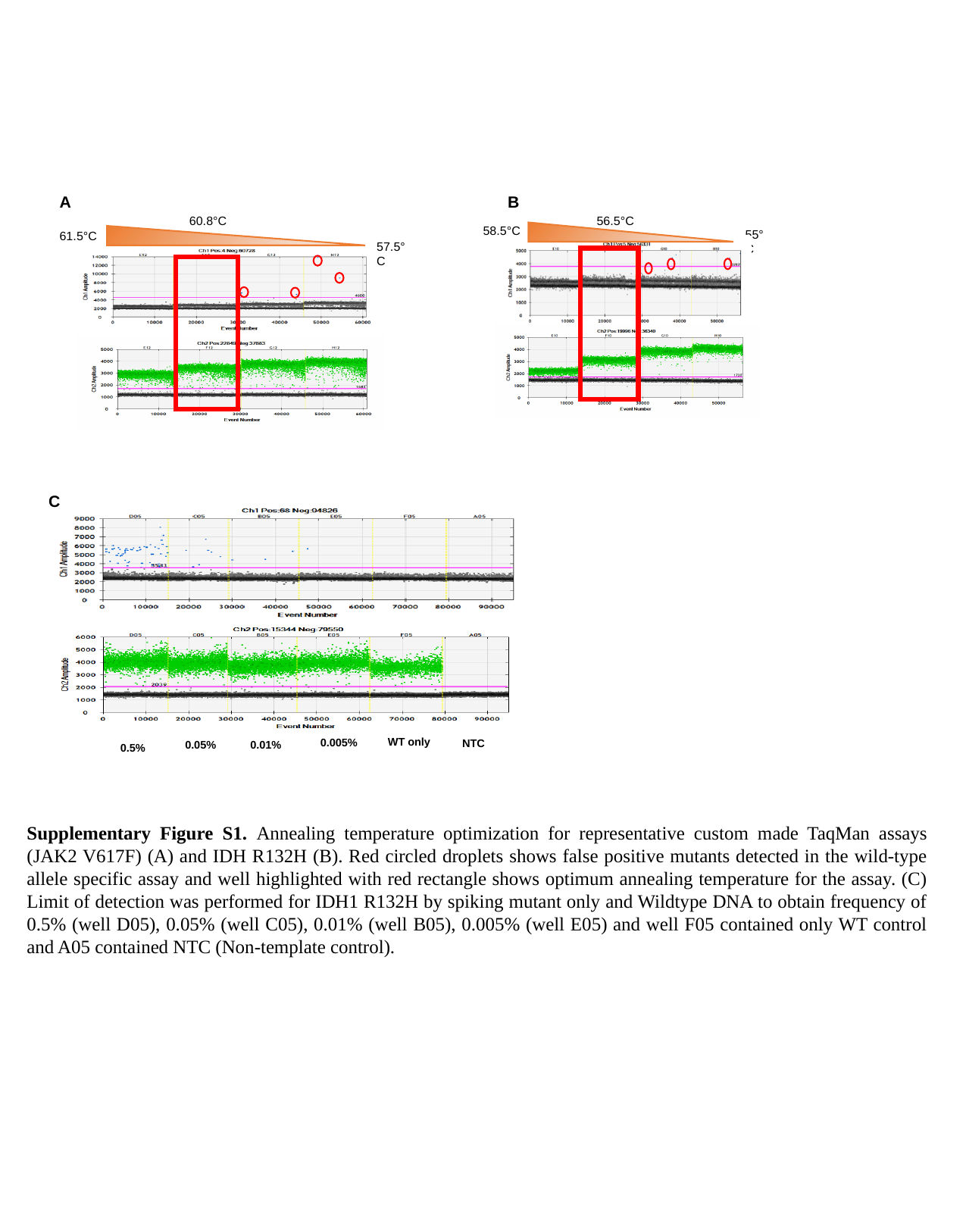

A
60.8°C
61.5°C
57.5°C
B
56.5°C
58.5°C
55°C
C
WT only
0.005%
NTC
0.01%
0.05%
0.5%
Supplementary Figure S1. Annealing temperature optimization for representative custom made TaqMan assays (JAK2 V617F) (A) and IDH R132H (B). Red circled droplets shows false positive mutants detected in the wild-type allele specific assay and well highlighted with red rectangle shows optimum annealing temperature for the assay. (C) Limit of detection was performed for IDH1 R132H by spiking mutant only and Wildtype DNA to obtain frequency of 0.5% (well D05), 0.05% (well C05), 0.01% (well B05), 0.005% (well E05) and well F05 contained only WT control and A05 contained NTC (Non-template control).

### Slide 2
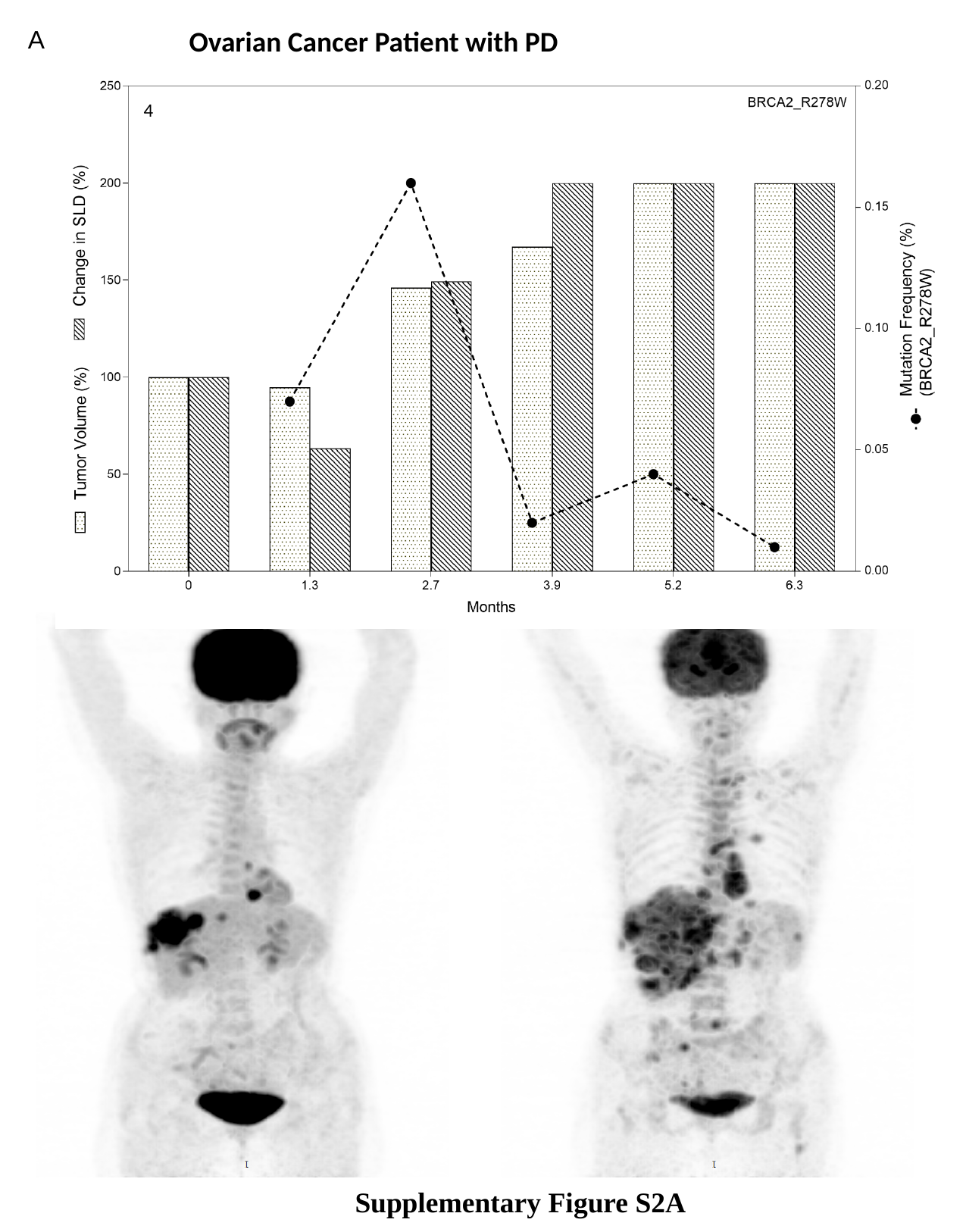

A
Ovarian Cancer Patient with PD
4
Supplementary Figure S2A

### Slide 3
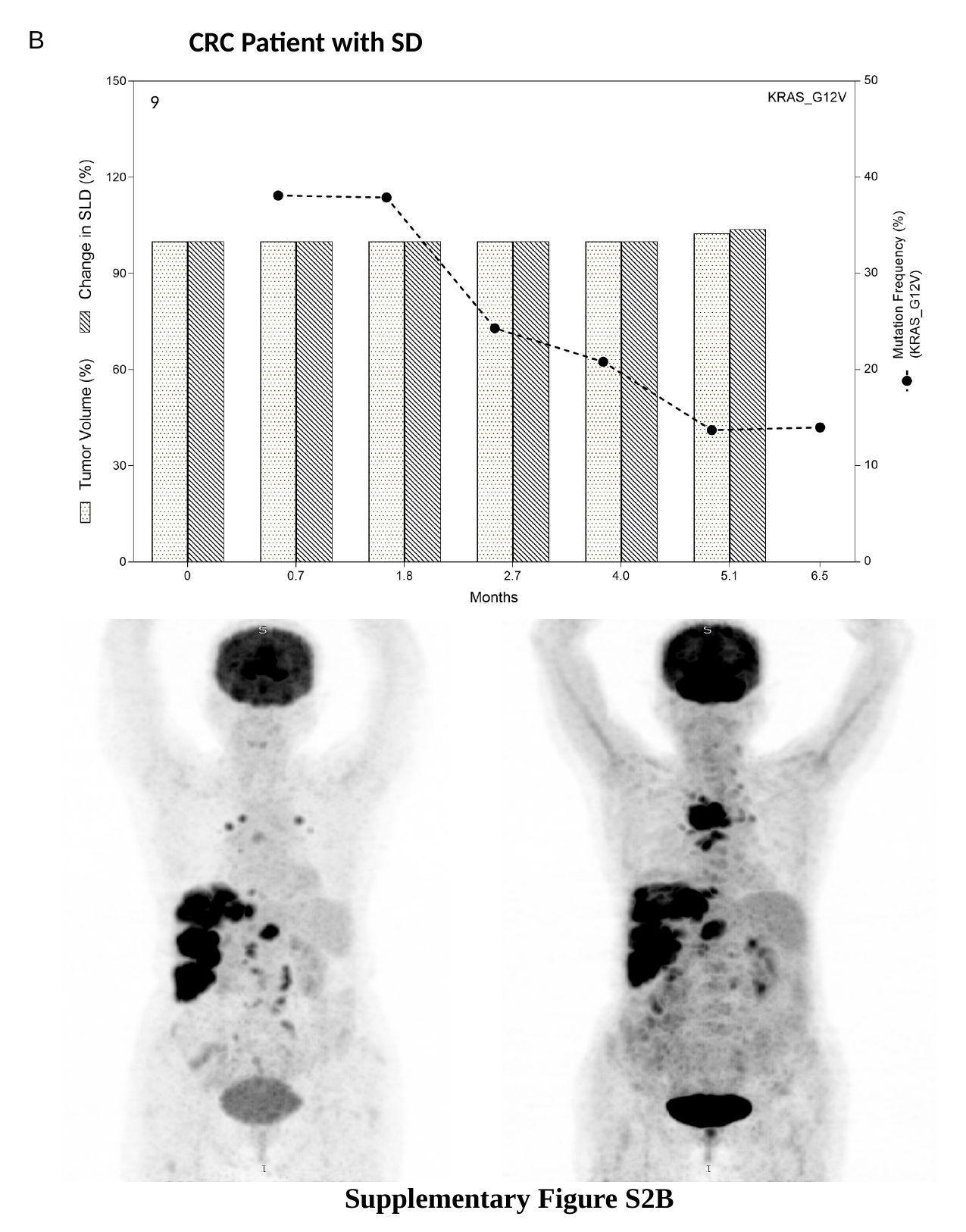

B
CRC Patient with SD
9
Supplementary Figure S2B

### Slide 4
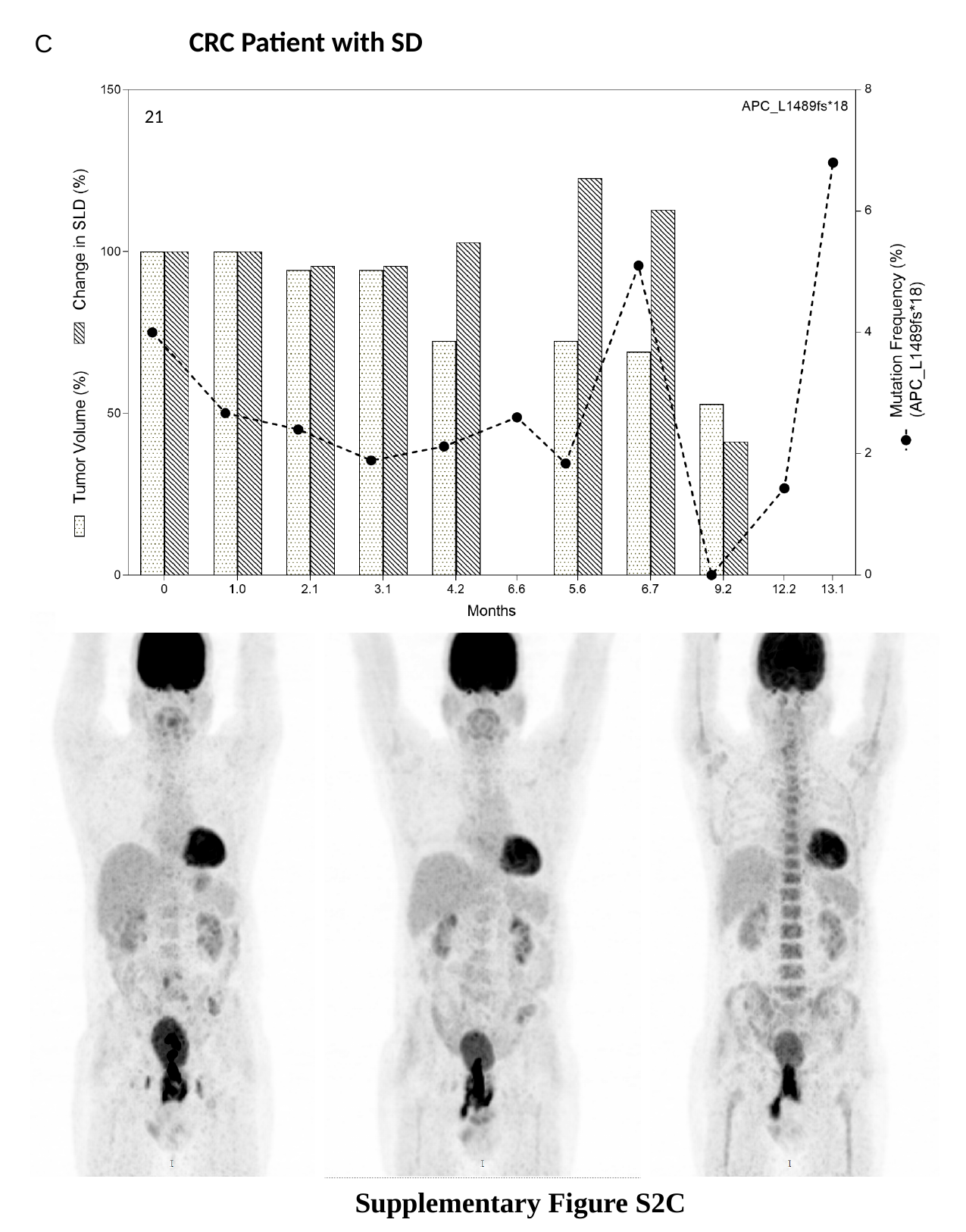

CRC Patient with SD
C
21
Supplementary Figure S2C

### Slide 5
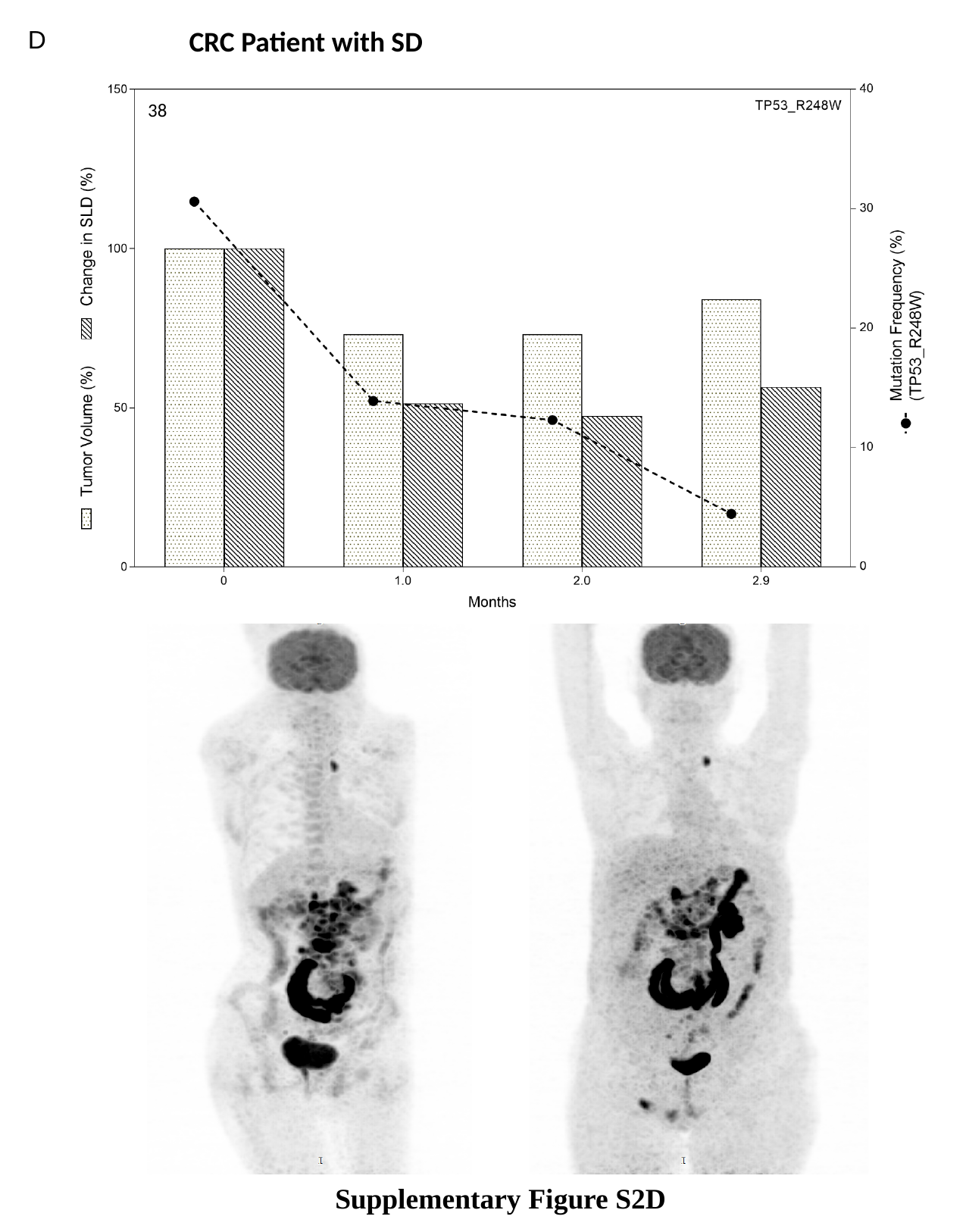

D
CRC Patient with SD
38
Supplementary Figure S2D

### Slide 6
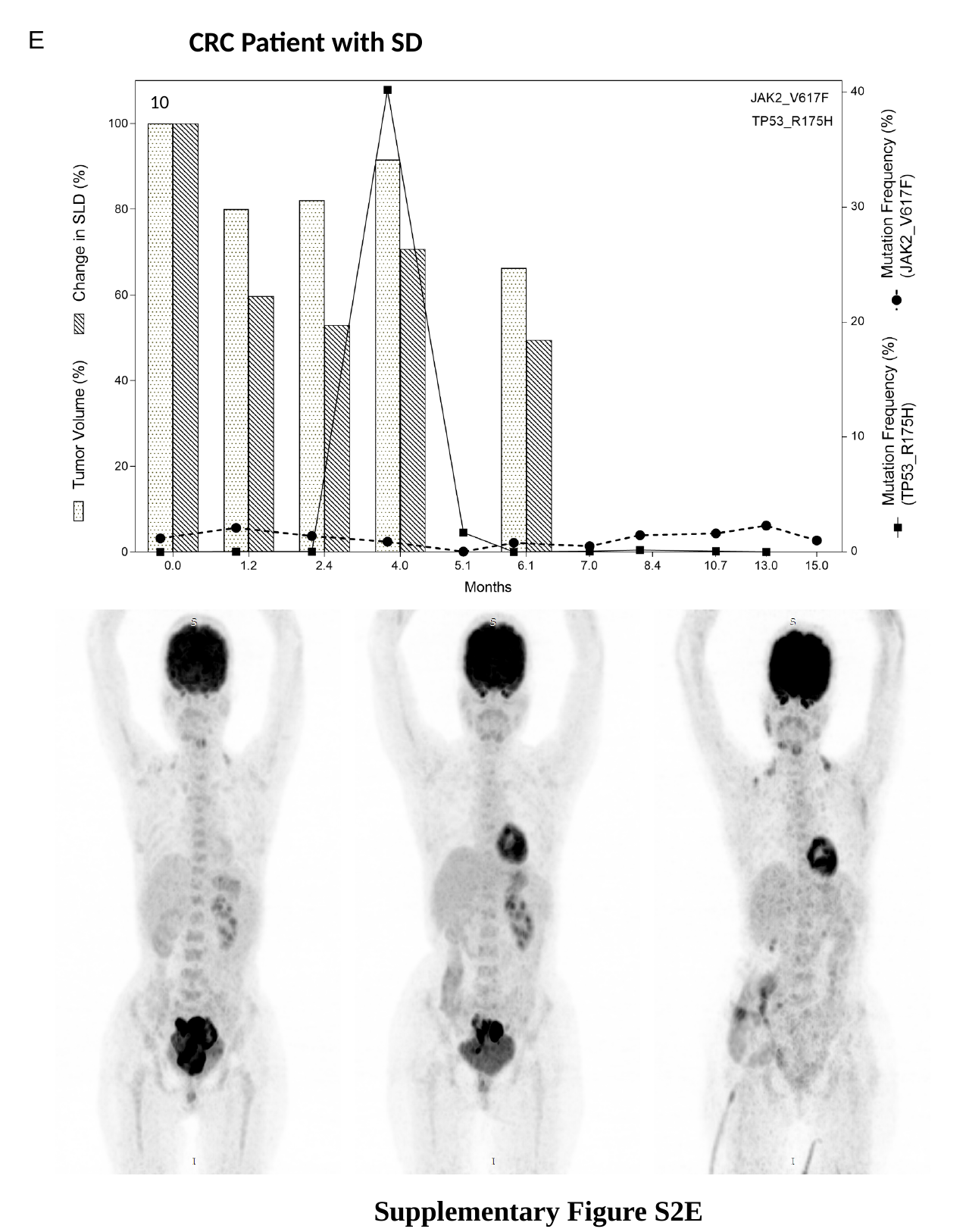

E
CRC Patient with SD
10
Supplementary Figure S2E

### Slide 7
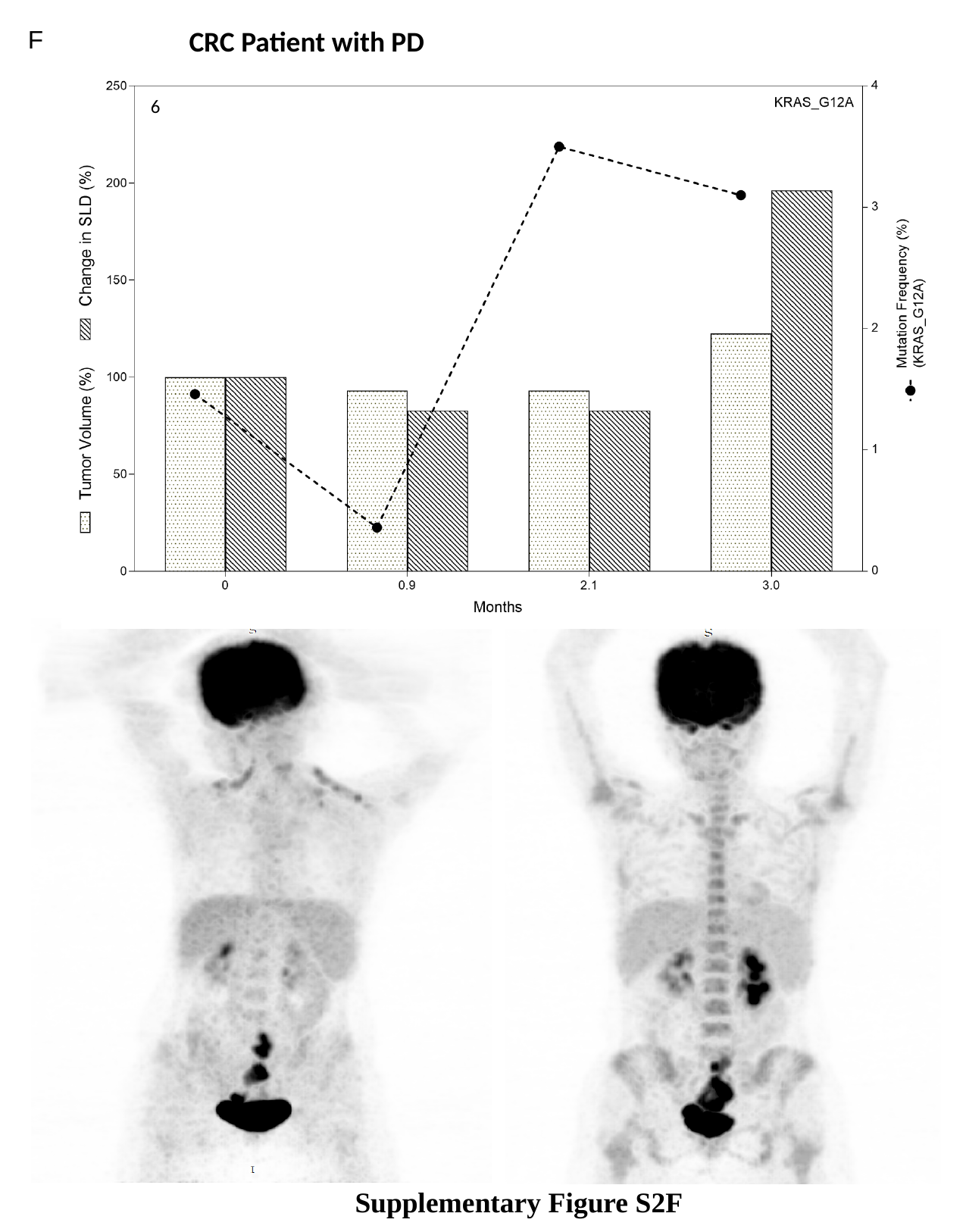

F
CRC Patient with PD
6
Supplementary Figure S2F

### Slide 8
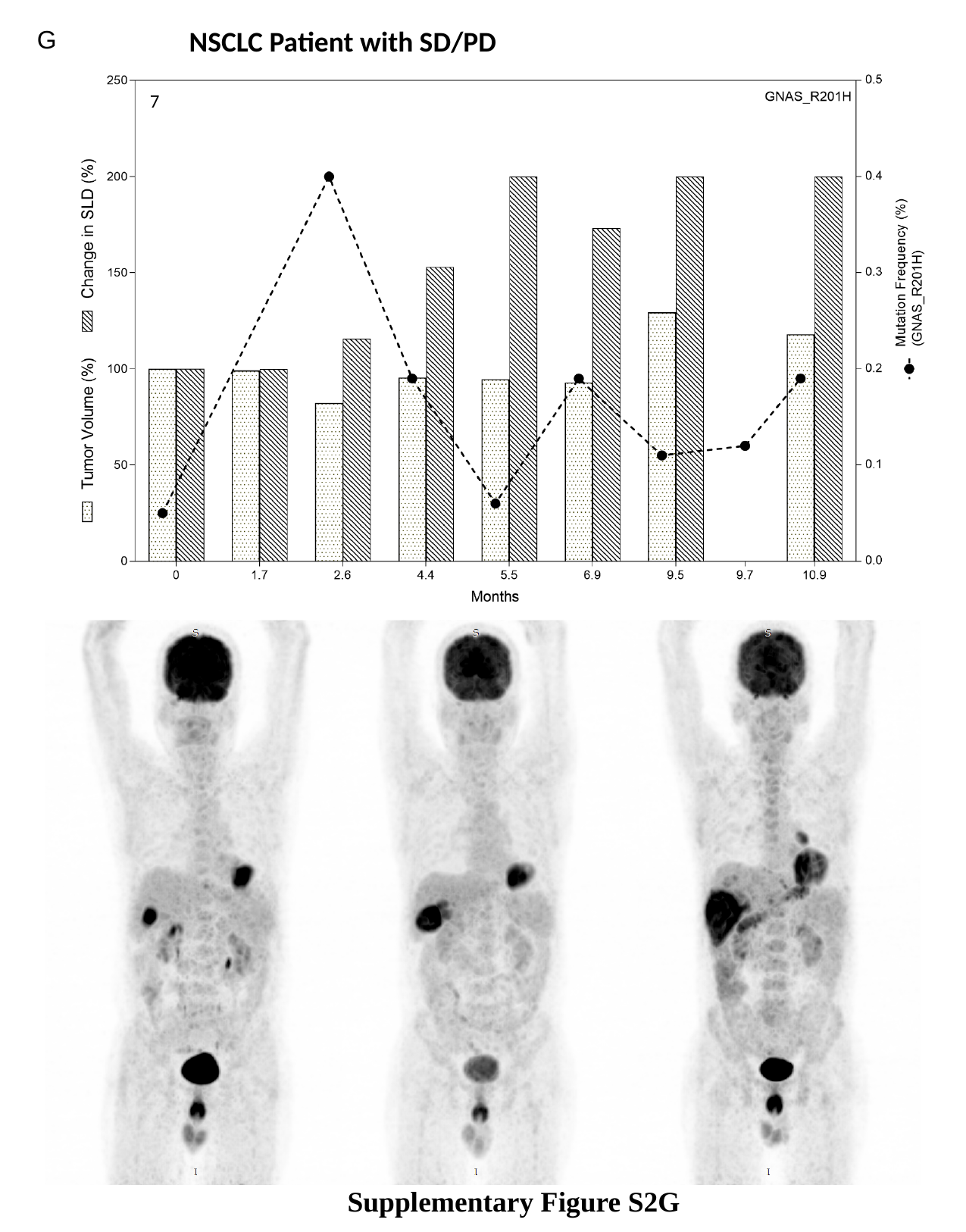

G
NSCLC Patient with SD/PD
7
Supplementary Figure S2G

### Slide 9
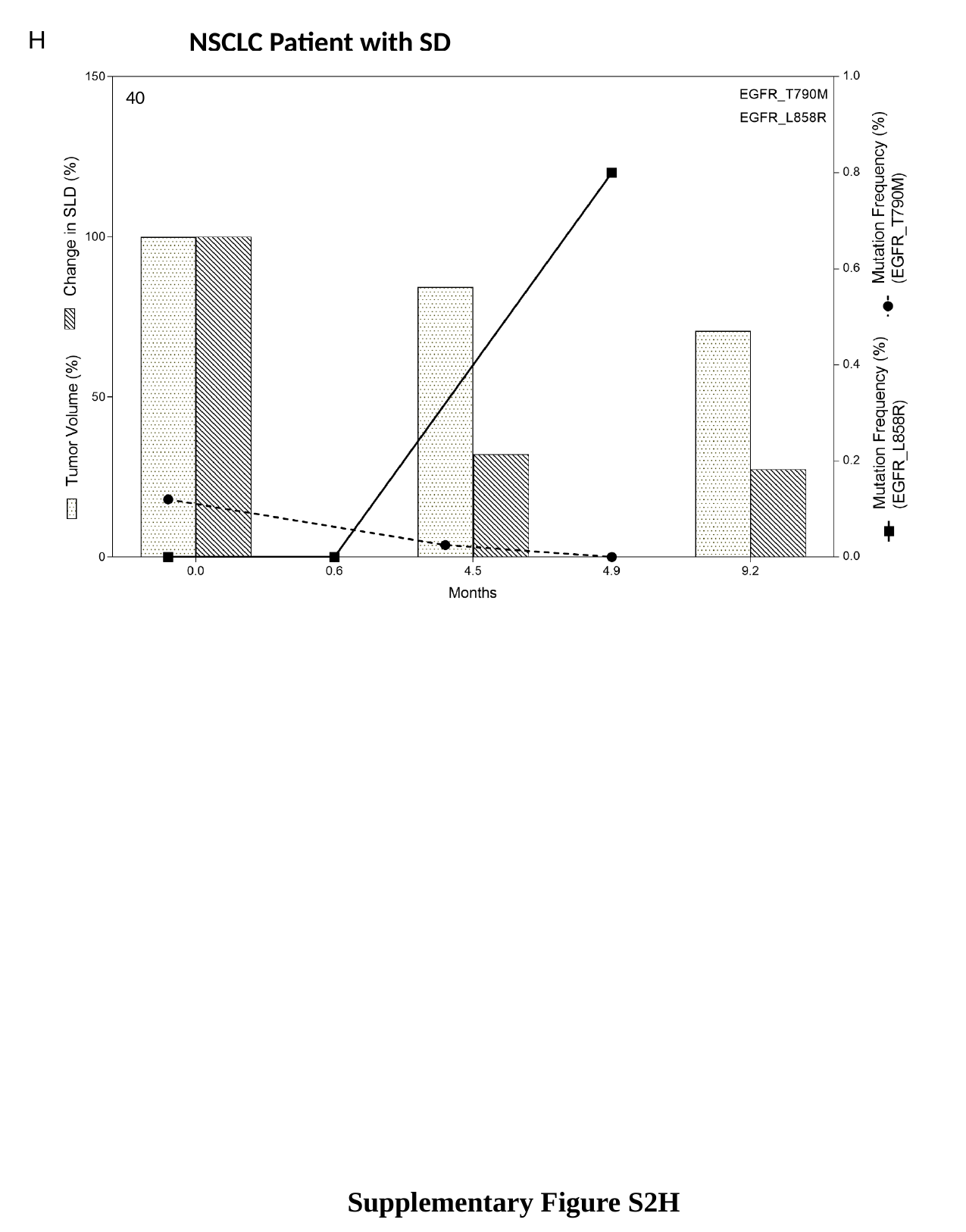

H
NSCLC Patient with SD
40
Supplementary Figure S2H

### Slide 10
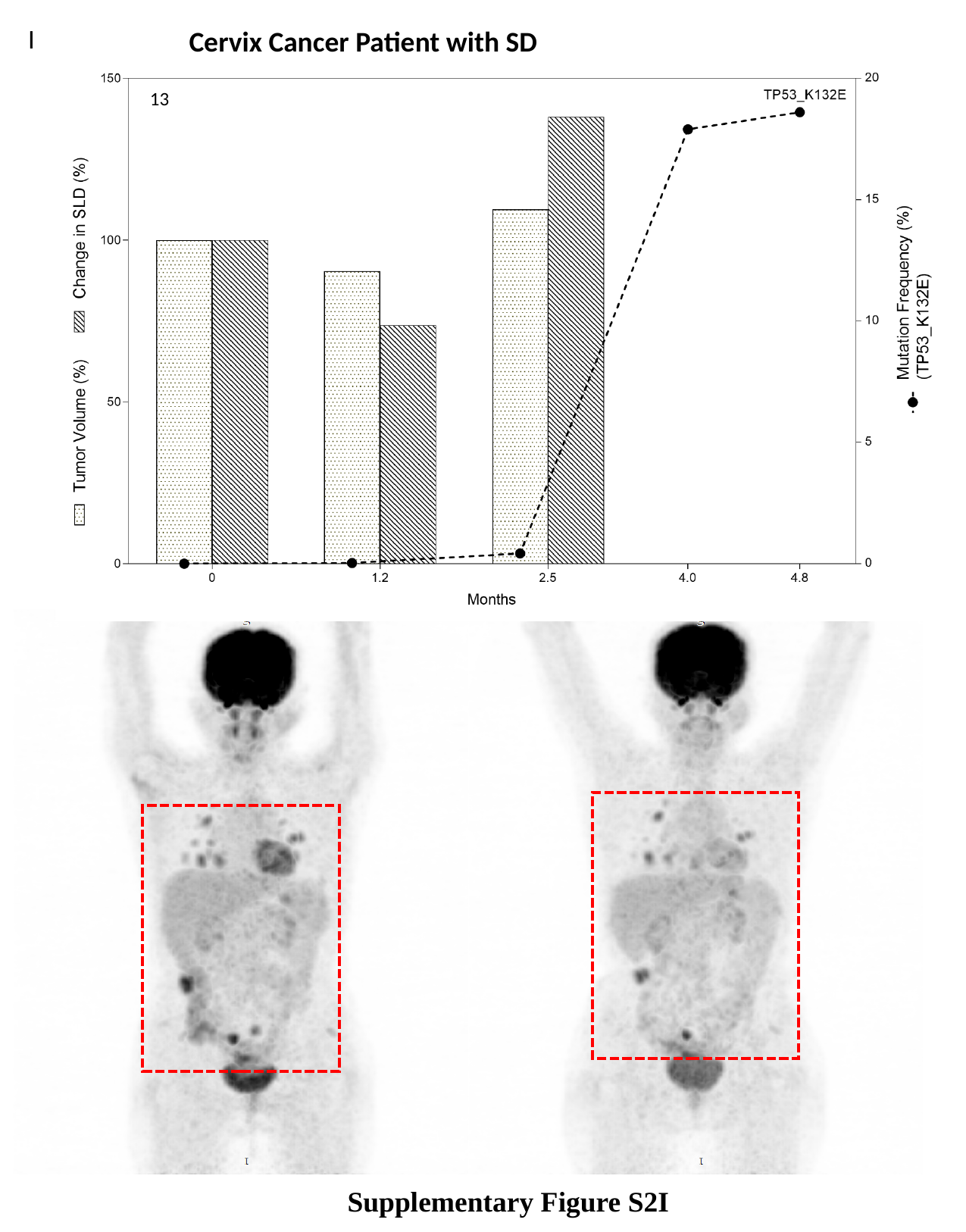

I
Cervix Cancer Patient with SD
13
Supplementary Figure S2I
